# COVID-19 outbreak in Greece has passed its rising inflection point and stepping into its peak

**DOI:** 10.1101/2020.04.15.20066712

**Authors:** Harris V. Georgiou

## Abstract

Since the beginning of 2020, COVID-19 is the most urgent and challenging task for the international scientific community, in order to identify its behaviour, track its progress and plan effective mitigation policies. In this study, Greece is the main focus for assessing the national outbreak and estimating the general trends and outlook of it. Multiple data analytics procedures, spectral decomposition and curve-fitting formulations are developed based on the data available at hand. Standard SIEQRDP epidemic modelling is applied for Greece and for the general region around it, providing hints for the outbreak progression in the mid- and long-term, for various infections under-reporting rates. The overall short-term outlook for Greece seems to be towards positive, with a downward trend in infections rate daily increase (i.e., now beyond the exponential growth rate), a possible peak within a few days beyond April 14th, as well as the high availability level of ICU w.r.t. expected demand at peak. On the negative side, the fade-out period seems to be in the order of several months, with high probability of recurrent surges of the outbreak. The mitigation policies for the ‘next day’ should be focused on close tracking of the epidemic via large-scale tests, strict border checking in international travelling and an adaptive plan for selective activation of mitigation measures when deemed necessary.

**Significance Statement:** This study focuses on the COVID-19 outbreak in Greece and provides data-driven epidemic modelling and experimental results regarding the current state. Based on these results, the overall short-term outlook for Greece seems to be towards positive, having recently passed the rising inflection point and approaching the peak of the infections, and most probably capable of covering the projected ICU demand peak by a large margin. On the downside, the fade-out period seems to be in the order of several months, with high probability of recurrent surges of the outbreak. The ‘next day’ mitigation policies need to be carefully planned, highly adaptive and based on close tracking of the outbreak via large-scale testing in the general population.

---

**B**acktrace of events and the availability of reliable data have enabled the construction of the timeline of events leading to the SARS-CoV-2 epidemic in China that has now turned into a world-wide pandemic within a few months. Based on data-driven modelling of the outbreak in various countries and cross-country regions, along with ‘synchronization’ to imposed policies and mitigation measures, it is possible to formulate a realistic outline of events, effects on the outbreak, as well as hints on how the situation will evolve in the next few weeks.

This paper is focused primarily on characterizing the SARS-CoV-2 outbreak in Greece, based on data as they become available on a daily basis from national and international sources. A short review of the timeline and the pathology of the virus are presented briefly; next, data analytics on the outbreak on the national level are providing hints and baseline parameters of the epidemic in Greece; the SEIR-like approach is introduced as a standard tool for modelling and predicting the outlook and the general outline of the next-day mitigation strategies; finally, a general discussion is made on how the pandemic evolves and how to address the challenges that lie ahead.

## 1. Short timeline of events

On December 31st 2019, when the western world was celebrating the New Year’s eve, China officially reported a cluster of cases of pneumonia-like illness of unknown cause in Wuhan, Hubei Province. The next day officials close the Huanan seafood market, suspected to be the source. A week later China identified a novel coronavirus as the cause of disease, which by then was already spreading in the local population. The first death by the disease, originally called ‘Wuhan pneumonia’ and officially labeled by WHO as ‘COVID-19’, was reported in Wuhan on January 9th 2020. Although the genome of the virus was readily available since January 12th, the virus continued to spread across the region and by January 20th its was already infecting people in Thailand, Japan and South Korea. The next day WHO confirmed human-to-human transmission of the virus, unofficially tagged as ‘2019-nCoV’ at that moment, while China was already reporting 580 confirmed cases. Beginning on January 23rd, the cities of Wuhan, Ezhou and Huanggang announce a general lockdown regarding public transportations, events and public gatherings.

By the end of January the virus had already reached Europe (Germany) and Australia, while China was reporting hundreds of new cases with a doubling rate of almost one day. Since most of non-Asian countries were still imposing no restrictions to international flights, it was clear by then that this was going to be a world-wide pandemic of unknown proportions. Indeed, by February 9th the international death toll of victims of the new virus was more than 800, surpassing the fatalities of the SARS epidemic in 2002 and 2003 that claimed 773 people. On February 14th Egypt reported its first confirmed case, the first African country in the recent 10 days, and on 15th France reported its first fatality from the virus, the first outside Asia. Two days later a paper from Chinese researchers provided some preliminary details about the virus, showing that COVID-19 is not as deadly as other types of coronaviruses, with about 80% of patients having mild disease, 14% severe disease including pneumonia, about 5% critical diseases including respiratory failure, septic shock and multi-organ failure and about 2% leading to death. These numbers were to be revised later on, but provided the general profile and pathology of the virus.

On February 19th the world-wide death toll from COVID-19 surpassed 2,000 including cases from more than 12 countries besides China. WHO continued to point out the severity of the spread and the lack of will and funding from the international community to address it properly and promptly. Israel and other countries in the region were already reporting confirmed cases by February 21st, a significant event for tracing the initialization of the epidemic in Greece about a week later on February 26th. Additionally, group of several dozens of people in total, tourists returning from Israel & Egypt and their close contacts, were quarantined after the first confirmed case was reported in northern Greece. About the same time, another group of Greek travellers had already returned back home from a business conference in northern Italy, while the border checks for the virus in the airports were still relatively relaxed (only some fever checking or none at all). This was the kick-off moment for Greece and at the time of new cases reporting being larger outside China than inside it.

The WHO officials had not yet declared a pandemic, but on February 28th raised the risk from ‘high’ to ‘very high’, stating that in such a case global travel restrictions would have a ‘significant economic and social impact’. In the following week the world-wide number of confirmed cases surpassed 100,000 and almost every country in Europe was now starting to deal with national outbreaks.

On March 11th the WHO officially declared the global COVID-19 outbreak as ‘pandemic’ and Greece goes into lockdown on schools, social events and public gatherings, following a few days of escalation towards measures of social distancing and self-isolation recommendations. On March 16th the international toll of COVID-19 fatalities outside China surpass those of inside China, the source of the outbreak. From there on, the exponential growth of the spread is ravaging Europe and the rest of the world, especially Italy, Spain, France and the USA, with no solid signs of slowing down, although some countries with containment measures (see: Figure 1) set early on seem to be in control and at the brink of a downward trend in newly reported cases.

**Fig. 1.**
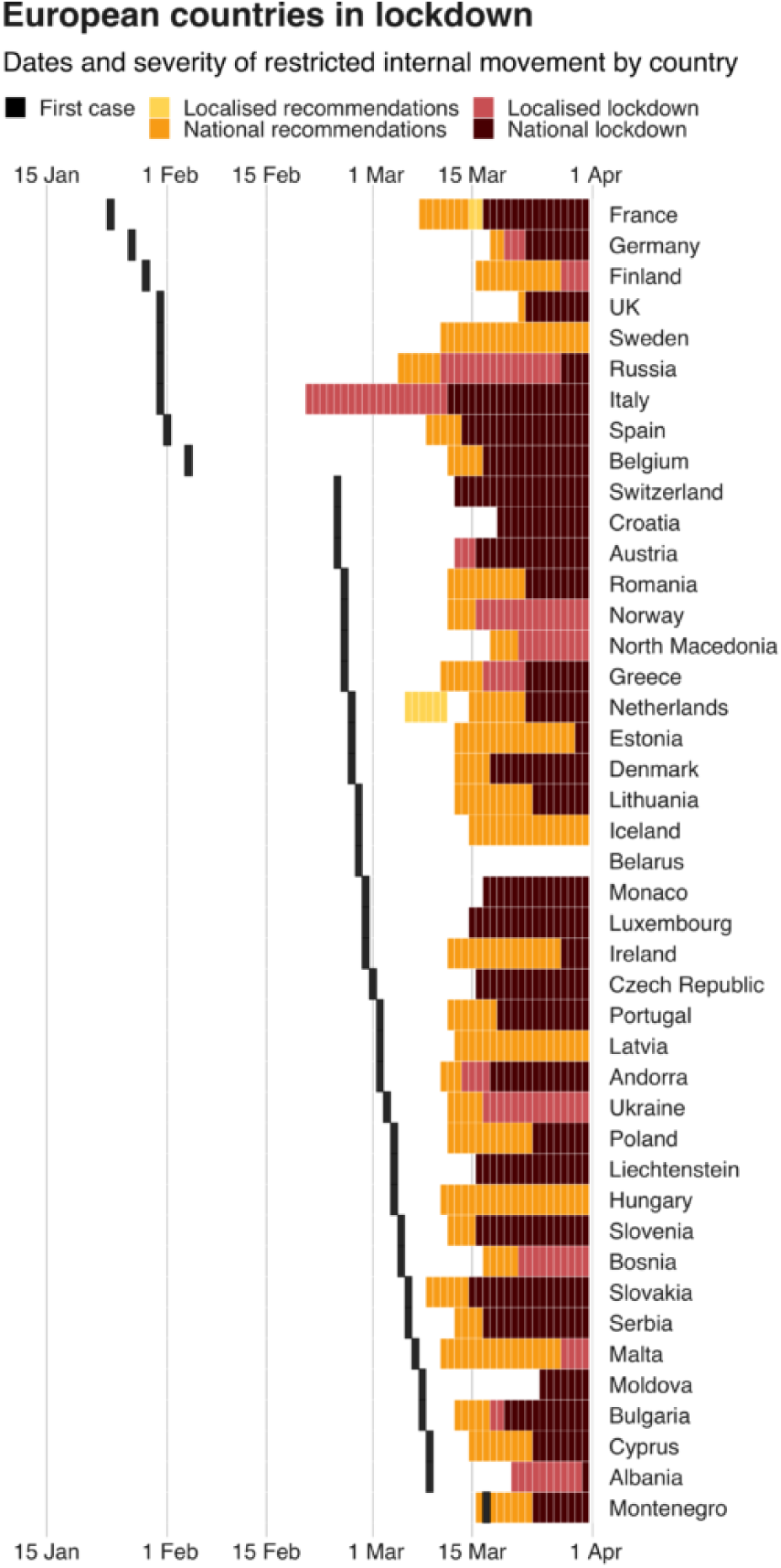
Per-country timeline of COVID-19 mitigation measures. (Source: Oxford Gov. Response Tracker / Credits: BBC)

### A. Outbreak onset and timeline in Greece

Greece seemed to have a slow start regarding its internal SARS-CoV-2 outbreak. By early February 2020 it was clear that the virus is going to land inside the borders at some point after a few days of a couple of weeks at most. Additionally, reports coming from other European countries already affected, as well as from China which still is the main source of information regarding the pandemic and the virus itself, gave time to the authorities to plan a gradual escalation of measures.

The timeline of the main events in Greece, followed by quick escalation of restrictive measures towards the final lockdown (Mar, 23) is as follows:

- Feb, 25: A few suspected cases reported in Patras, travellers on a ship from Italy.
- Feb, 26: 1st confirmed case reported, a woman who returned from northern Italy.
- Mar, 04: 1st confirmed case in the group of tourists who returned from Israel & Egypt.
- Mar, 05: Case-based self-isolation mandated by the authorities.
- Mar, 06: Most of the tourists who returned from Israel & Egypt were confirmed as infected.
- Mar, 07: Social distancing ‘encouraged’; public venues & events cancelled in three cities.
- Mar, 08: 1st confirmed case of unknown origin; detailed tracing of contacts is now infeasible.
- Mar, 09: Four more confirmed cases of unknown origin; all public events cancelled/banned.
- Mar, 11: All schools ordered to close ‘for at least two weeks’.
- Mar, 12: 10 new confirmed cases of unknown origin; 1st virus-associated death; universities close.
- Mar, 19: 464 confirmed cases in total; hotels & venues close.
- Mar, 20: Reports of increased traffic away from large cities.
- Mar, 23: Country-wide lockdown in effect.

Besides the first confirmed case on February 26th, the first important milestone in this timeline is the 72 hours between March 4th-6th, when the entire group of tourists who returned from Israel and Egypt (both already in national outbreak since February 21st and 14th, respectively) was tested and most of them were confirmed as infected. The next milestone is on March 8th, when the first confirmed case of unknown origin was reported. The next day this number was four and four days later it was 10, which translates to a spreading rate of 1.778 < *R*_0_ *<* 4 for these cases of ‘unknown origin’. From there on, it was almost certain that it was now infeasible anymore to contain them in time with backtracing and targeted isolation. The third and most vivid milestone, i.e., the country-wide lockdown measures, was an already expected outcome by the public since the 2-3 previous days, as the outbreak in Italy was starting to explode.

Figure 2 illustrates the confirmed cases of infections in Greece per region on March 15th, a week before the country-wide lockdown went into effect and while the outbreak was starting to significantly increase speed.

**Fig. 2.**
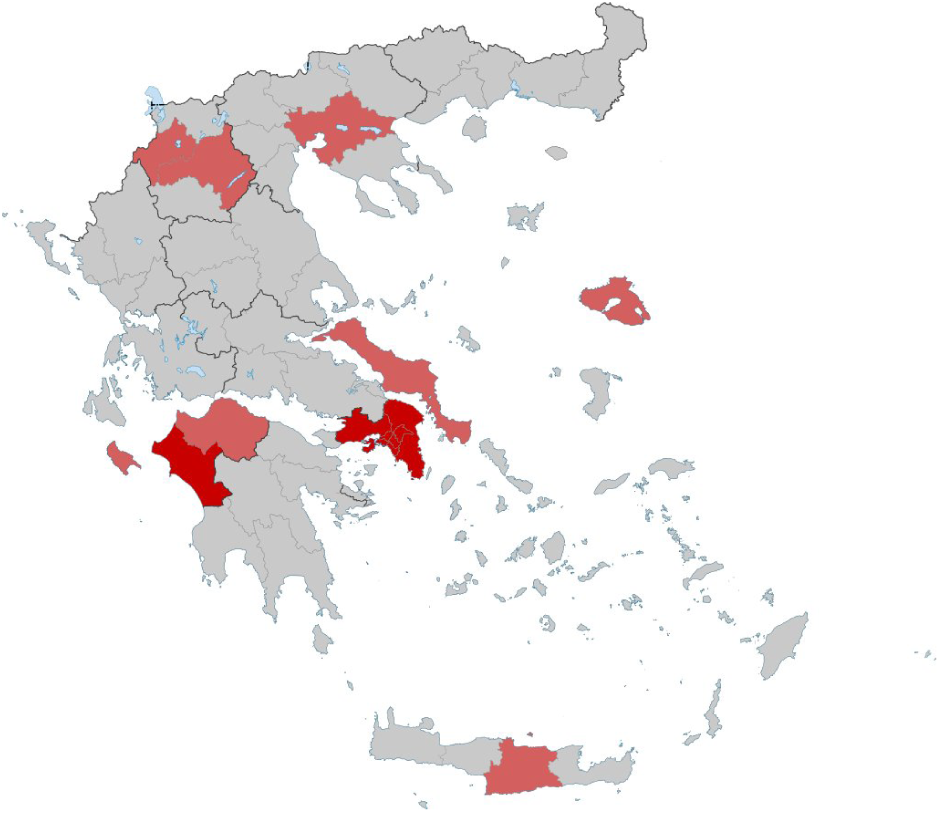
COVID-19 heatmap of confirmed cases of infections in Greece on March 15th, 2020. (Source: Wikipedia.org)

It is worth noting that, although since March 23rd Greece is in lockdown, some businesses are still allowed to operate; these are primarily pharmacies, supermarkets, banks, groceries, bakeries, etc (food, medicines, basic needs). Movements of individuals is allowed only to/from work and to/from home for limited trips of shopping, pet walking, athletics (single). No public venues or events are allowed at least up to the middle of May. There are strict checks at the tolls and penalties for travellers and even individuals without carrying proper paperwork. For still-working persons, driving in a personal car is encouraged and public transport, although still functioning, has severely limited the scheduled trips per route.

## 2. Characteristics of COVID-19

Coronaviruses constitute a large family of viruses that can cause a wide range of disease cases, ranging from common colds to severe pneumonia and fatalities, usually from secondary cases and other underlying pathologies. They are usually found in animals and some are transmittable to humans, which may later mutate and become transmittable between humans too. SARS and MERS diseases are caused by a corresponding coronavirus and their pathological characteristics are well-studied in the last two decades.

Early in the epidemic in China, the SARS-CoV-2 virus was isolated and its genome analyzed. Figures 3 and 4 illustrate two of the very first pictures of the virus via electron-microscope. It was quickly identified as a SARS-like in type, pathology and estimated epidemic characteristics. Soon after the first few hundred of confirmed cases in China were treated in hospitals, a general profile of its symptoms started to emerge and the doctors throughout the world could immediately identify a ‘possibly positive’, as Figure 5 shows.

**Fig. 3.**
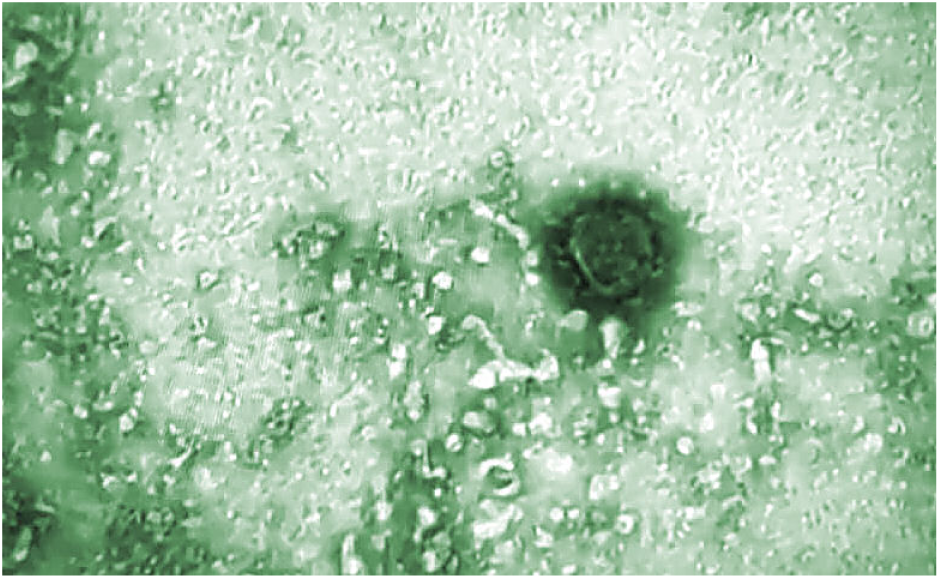
The first SARS-CoV-2 virus image via electron microscope. (Source: Greek media)

**Fig. 4.**
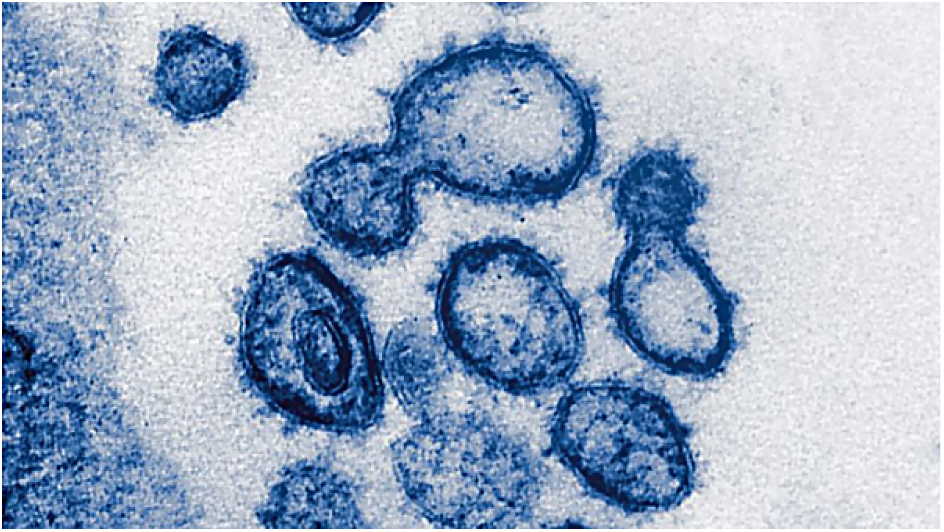
The SARS-CoV-2 virus by an electron microscope. (Source: National Institutes of Health - RML/SPL)

**Fig. 5.**
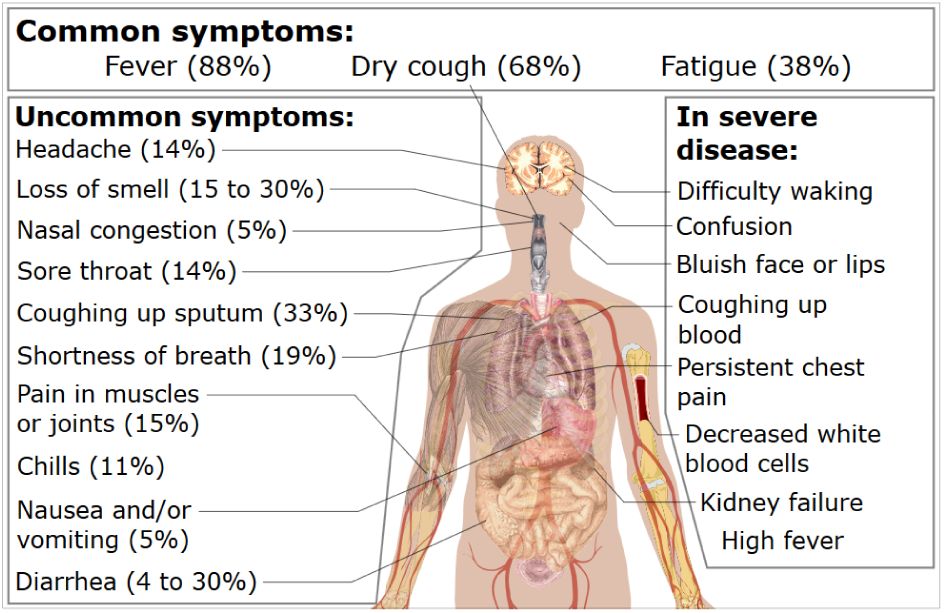
Common symptoms and effects of the COVID-19. (Source: Wikipedia.org)

According to the currently available world-wide data, the virus’ reproduction and contagious characteristics place it somewhere between SARS and Ebola (see: Figure 6). Its fatality rate is somewhat more difficult to estimate, as this depends largely on each country’s health infrastructure and capacity, the stage of the outbreak, if these facilities and saturated or not, other underlying pathological issues in the general population, etc. On April 14th (2020) the top WHO official Tedros Adhanom Ghebreyesus stated^*^ that COVID-19 is ‘ten times deadlier’ than the H1N1 strain (‘swine flue’), which evolved world-wide between January 2009 and August 2010, with more than 1.6 million confirmed cases and 18,449 deaths. He also added that in some countries the doubling period is ‘three to four days’ and that this virus ‘accelerates very fast it decelerates much more slowly’.

**Fig. 6.**
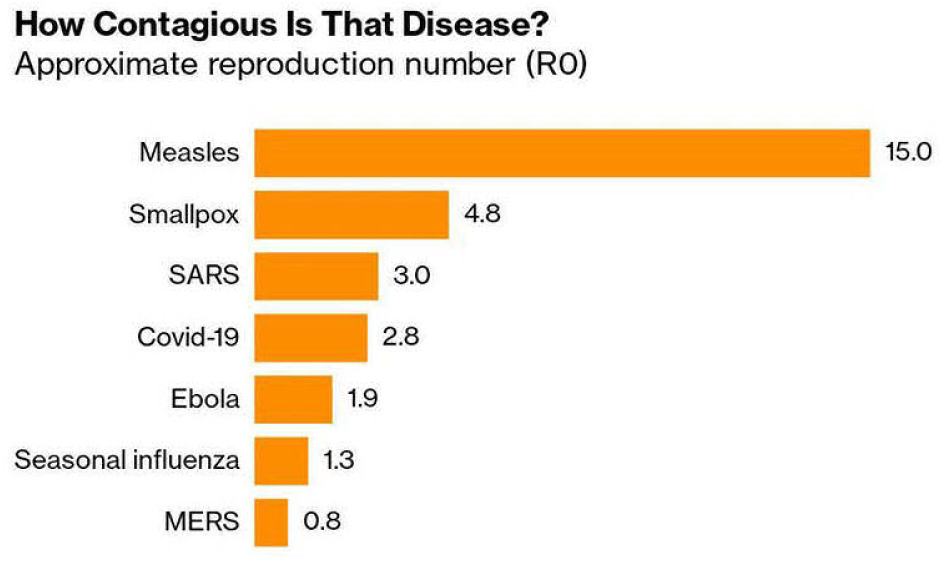
Estimated contagiousness of SARS-CoV-2, Mar. 2020. (Source: WHO, CDC, King Saud Univ., Nature / Credits: Bloomberg)

A first estimation places its fatality rate in China (early outbreak) at least above 3.4% of confirmed cases; however, post-analysis with extensive cases analysis, as well as results from countries with a very large rate of randomized tests (including South Korea, Iceland, Germany) lower the actual fatality rate to 1% or less (see: Figure 7). Taking into account the epidemic data from several countries within this first three-month window, Figure 8 shows the real range of the fatality rate, ranging from 0.25% to 10% or more, depending on the circumstances and phase of the epidemic.

**Fig. 7.**
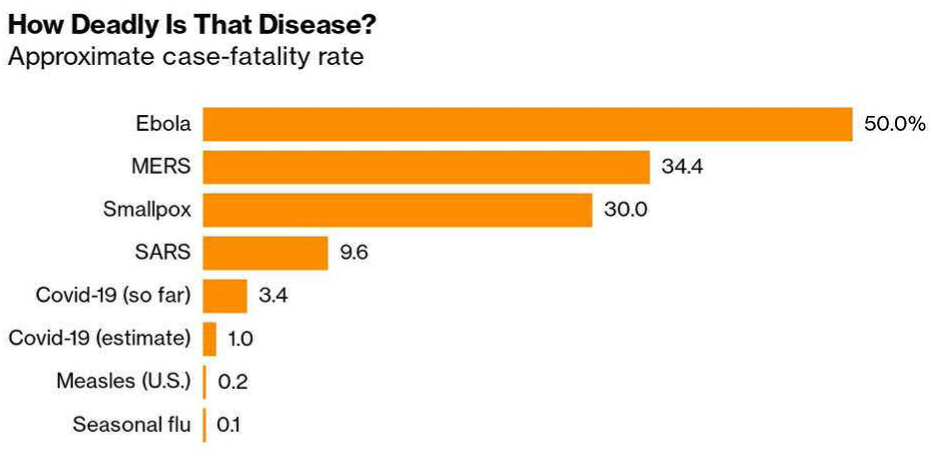
Estimated fatality rate of SARS-CoV-2, Mar. 2020. (Source: WHO, CDC, MRC Center for Global Infectious Disease Analysis / Credits: Bloomberg)

**Fig. 8.**
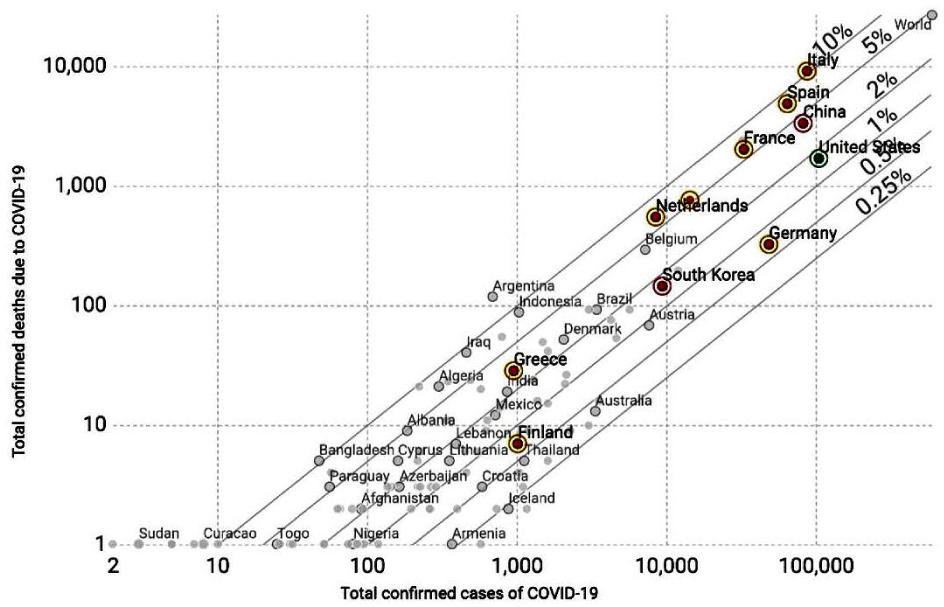
COVID-19 fatality rate per country, Mar. 2020. (Source: European CDC / Credits: OurWorldInData.org)

Overall, the world-wide pandemic seems to be evolving in exponential or sub-exponential rate depending on the country, time since the first (or first 100th) confirmed case and, primarily, the time of the onset of strict lockdown measures. Figure 9 presents the progress of the COVID-19 pandemic in various countries and continents, with higher curvature towards the right associated to slower spreading rates.

**Fig. 9.**
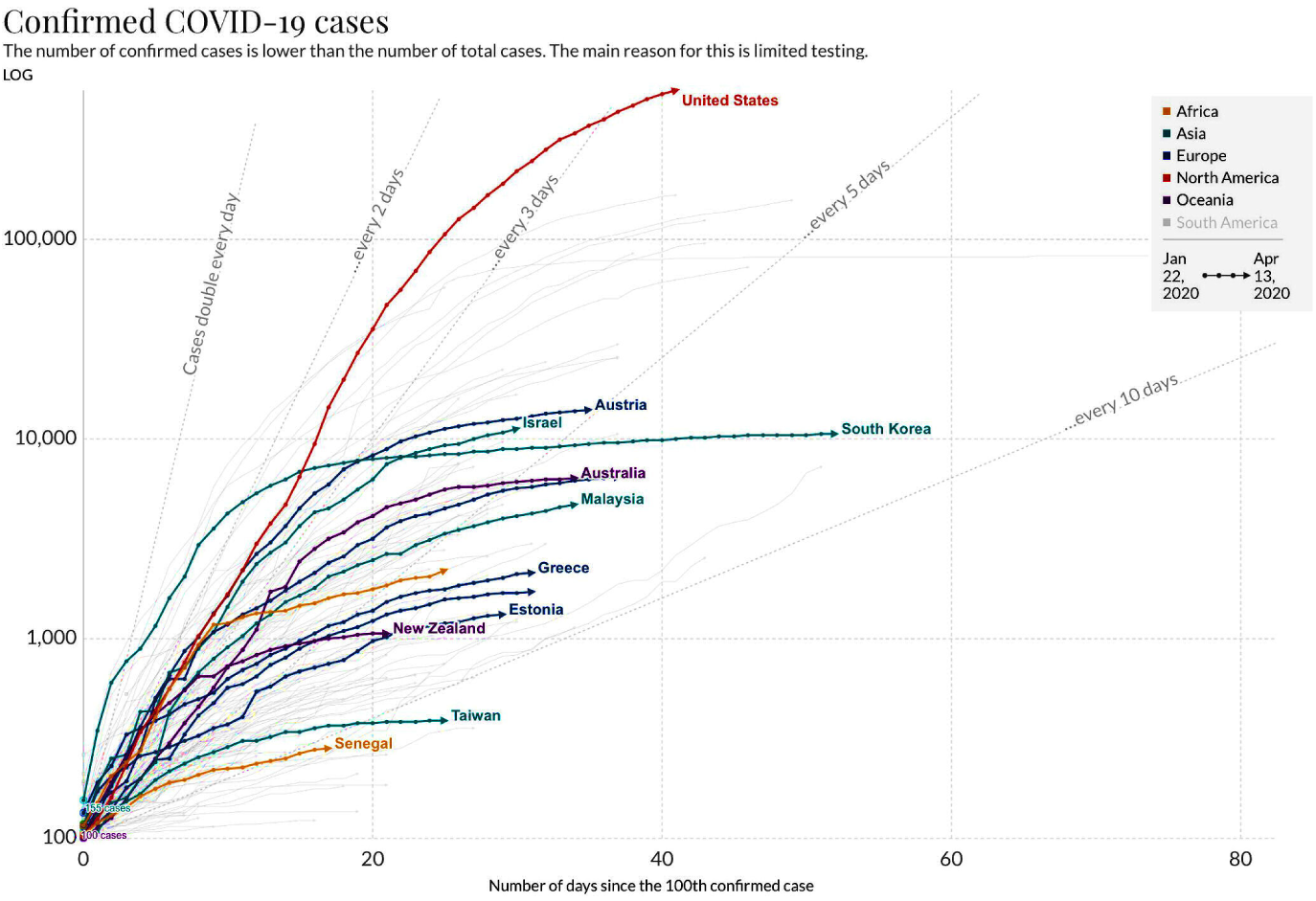
Progress of the COVID-19 pandemic in various countries and continents (logarithmic scale); higher curvarture towards the right is associated to slower spreading rates. (Source: European CDC / Credits: OurWorldIn-Data.org)

## 3. Epidemics

A virus outbreak in the general population of a town, a region or an entire country is a very dynamic and usually a rapidly evolving natural phenomenon. Depending on the speed of infections spread, the severity of the resulting diseases and the capacity of the local/national health system, this can be categorized as a natural disaster that may be very difficult to contain and mitigate.

There are several phases in a SARS-like virus epidemic, as Figure 10 illustrates. Phases 1-3 are mostly ‘dormant’, when the virus is mutated and begins to transmit form animals to humans, but remained undetected until the number of infections becomes significant. In phase 4 the virus is spreading freely in human populations, with the authorities not yet knowing how/where to address it and not yet having strict measures in effect to mitigate it. During this phase, the infections increase at a high exponential rate and a large portion of them is usually undetected and, thus, under-reported. Phase 5 may be split into two sub-phases: the first (phase 5a) begins at the moment the quarantine measures are in effect and start to gradually slow down the exponential growth to a peak (maximum number of active infections); the second (phase 5b) begins at the peak and continues to the point when the active infections decrease and the evolution is becoming a curve that asymptotically approaches zero. Then, phase 6 is the period of this asymptotically decreasing active infections, where usually the quarantine measures are gradually deactivated; this is the phase when most recurrent events occur, as some still-infected and unregistered cases can move freely again in the general population. Without prompt and effective vaccination procedures and large-scale tests in place for the general population, these events are almost certain to happen with flu-like viruses. After any such recurrent events fade out, the final phase is in effect and the virus spread can be tracked on a seasonal level, taking into account natural immunization (recovered patients) and vaccination that make a new large-scale outbreak very unlikely.

**Fig. 10.**
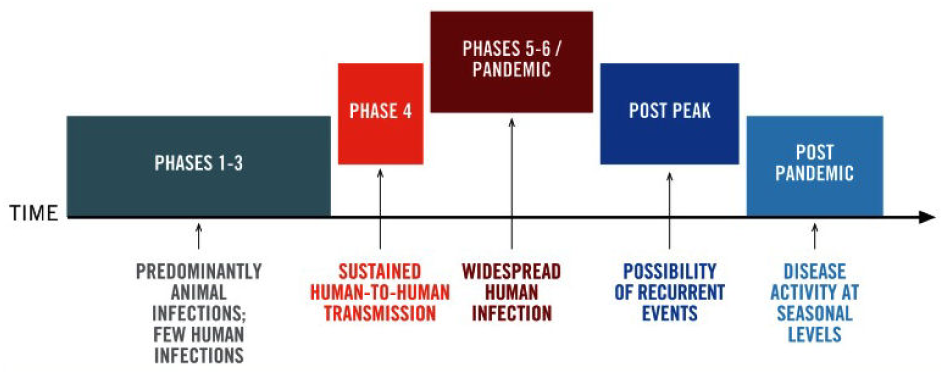
Typical phases of an epidemic outbreak. (Source: WHO)

Since early detection and stopping is only rarely a realistic option, in most cases the most critical phase in terms of mitigation is the middle or ‘peak’ of active infections (phase 5). Previous experience has proved that the most crucial factor there is the capacity of the health system that handles the concurrent active infections and most importantly: (a) the number of ICU available for critical cases, (b) the number of health workers that get infected or quarantined themselves, i.e., out of the ‘battle’, and (c) the availability of targeted tests and other resources, e.g., personal protection gear, spare parts for medical equipment, consumables in hospitals, etc.

In order to keep the infection rate within the capacity of the health system, i.e., below a critical threshold, it is imperative that social distancing and protective measures are early in phase 4 - this causes the infection rate to slow down and the peak to become lower, an effect that is often referred to as ‘flattening the curve’ (see: Figure 11). On the other hand, slowing down the infection rate results in much longer ‘tails’ in phase 6, i.e., a similarly slower rate of the asymptotically fading-out of the outbreak. Additionally, If no vaccination is available in phase 6, any deactivation of the quarantine measures re-exposes the general population to undetected infections and the probability of a recurrent outbreak becomes higher. In other words, the more ‘flat’ the curve is at the peak of the outbreak, the longer is the fade-out period is and with higher possibility of another outbreak. Additionally, since the infection-to-recovery cycle usually produces natural immunization (depending on the virus type), having a ‘flat’ curve also translates to lower levels of natural protection for the general public and usually stronger subsequent waves of the same outbreak. Figure 12 illustrates this effect with the ‘Spanish flu’ virus and how the outbreak evolved in Philadelphia, San Francisco, New York and St. Louis in 1918-1919.

**Fig. 11.**
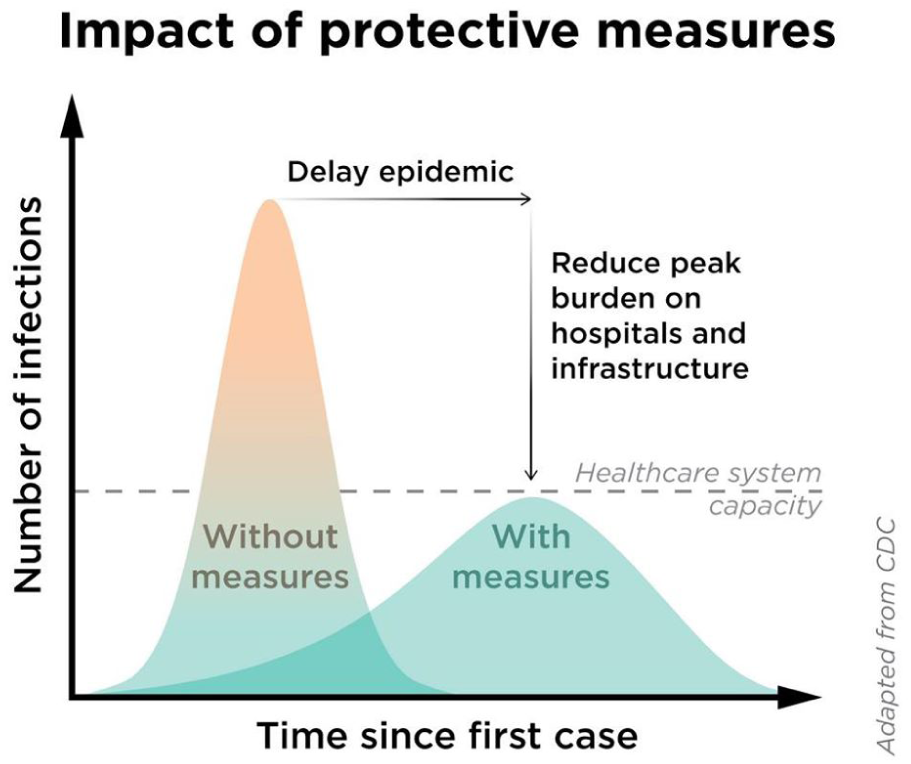
Infectios rate progress without and with ‘flattening the curve’ mitigation measures. (Source: CDC)

**Fig. 12.**
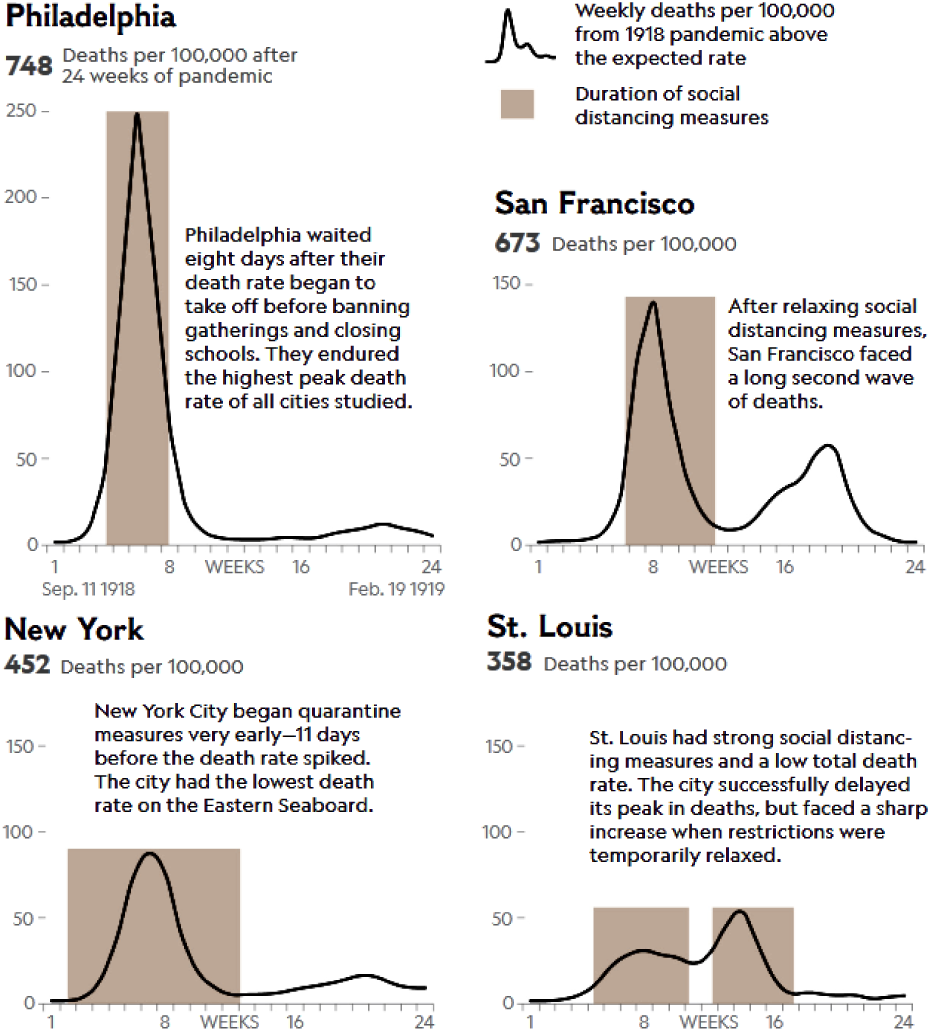
Infections rate and recurrent waves during the ‘Spanish flu’ outbreak in four major US cities, 1918-1919. (Source: National Geographic)

In the end, if the complete wipe-out of the virus cannot be guaranteed in the long term, the only stable outcome is when the general population has been immunized, naturally (infected/recovered) or artificially (vaccination), at a minimum proportion usually much higher that 60-70%, depending on the type of the virus - but still, 100% is not required, due to the ‘herd immunity’ phenomenon (1, 2). Hence, ‘flattening the curve’ is a measure of securing more time for the health system to cope with the highest infection rate or ‘peak’ and for the community to improve intermediate treatments and develop a vaccine for large-scale production.

## 4. Compartmentalized SIR/SEIR models

The mathematical modelling of epidemics has been a very active research field for decades, even before sources with detailed data were available. As explained in section 3, it is important to be able to estimate the progress of the outbreak and its phase, in order to properly and promptly plan the mitigation measures.

By far, the most popular and well-established approach is the family of *compartmental epidemic models*, originally developed as far back as 1920s. Their common characteristic is the base assumption of having a target population partitioned in *compartments* that are homogeneous in all relevant properties (e.g., sex, age, underlying pathologies, etc) and there are direct interactions between them. The three basic compartments are S=‘susceptible’, I=‘infectious’ and R=‘recovered’, assuming insignificant rate of deaths and permanent immunity after recovery. Variants of this SIR base model include a D=‘deaths’ compartment (SIRD), E=‘exposed’ compartment (SEIR, SEIRD) for introducing an incubation period, Q=‘quarantined’ compartment (SEIQRD, SEIQRDP) for separating the already isolated confirmed/possible carriers, etc. The SIR-like models, along with more recent SEIR-like variants, are still used as the baseline for comparing other approaches to epidemic modelling. These are usually based on Sequential Monte Carlo (3, 4), Markov Chain Monte Carlo (MCMC) or Markov Chain quasi-Monte Carlo (MCQMC) (5– 7), Hidden Markov Models (HMM) (8–11), etc, each posing other assumptions, advantages and drawbacks - most commonly the availability or not of a significant amount of epidemic data upon which they are to be trained.

The SIR-like epidemic modelling is closely related to the Lotka–Volterra system of equations (12), developed in the 1910s to describe the evolution of dynamic systems via differential equations. They constitute an example of the more generic Kolmogorov model (13, 14) which can describe the dynamics of ecological systems with predator–prey interactions, competition, disease, etc.

Based on recent models that are already being tested with COVID-19 data from China and other countries, the current work explored the (still scarce) epidemic data for Greece via the generic framework of a SEIQRDP model setup (15, 16).

The additional P=‘insusceptible’ corresponds to a fraction of the general population (if any) that, even when exposed to the virus, cannot become ‘infected’ and, thus, does not enter the E compartments and stays outside the ‘pipeline’ of the epidemic. Each interaction between the SEIQRDP compartments is governed by an scalar parameter that governs the way fractions of each subset is ‘transferred’ to another, e.g., from ‘infected’ to ‘recovered’. Figure 13 illustrates the SEIQRDP model and the meaning of each parameter. The internal structure of the model, i.e., the interactions that describe the dynamics of the system, is formulated by Eq.1 through Eq.7.

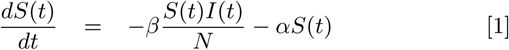

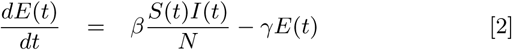

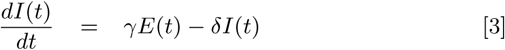

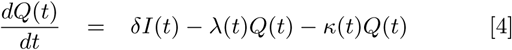

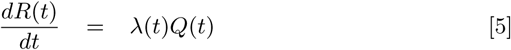

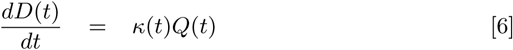

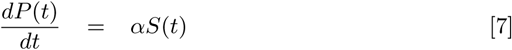

**Fig. 13.**
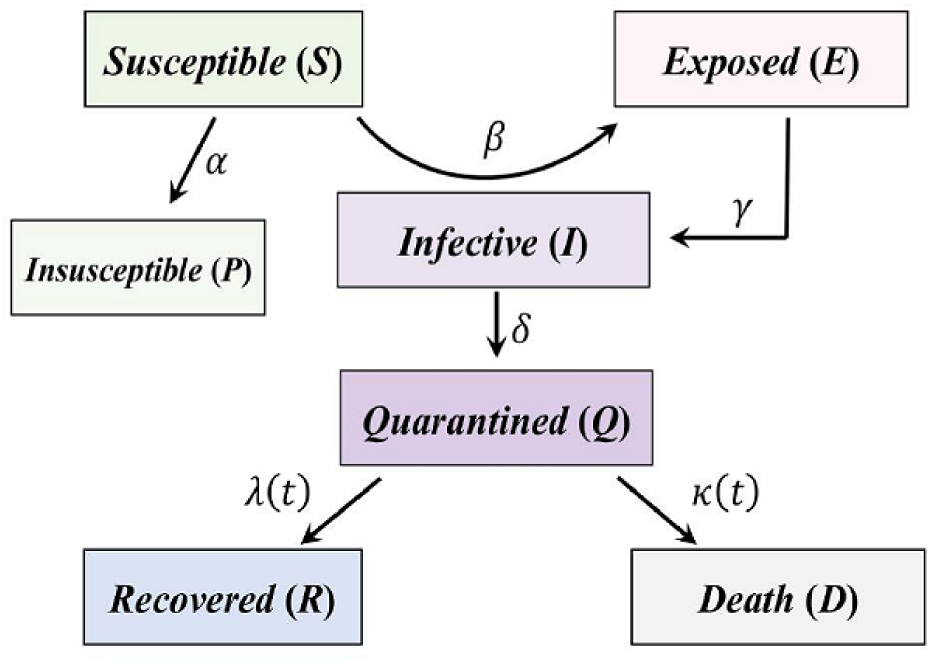
The block diagram of the SEIQRDP model and its parameters.

The system of SEIQRDP model are typical first-order linear differential equations (17) in the form of *dy/dx* + *P* (*x*)*y* = *Q*(*x*), which can be solved analytically according to Eq.8:

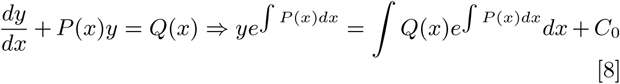

However, these analytical solutions are useful mostly for estimating the asymptotic steady-state outcomes and only when the equations are fully defined, i.e., w hen a ll the function parameters are already known (18). For example, Eq.1 in SEIQRDP can be analytically solved according to Eq.9 and Eq.10, i.e., by substituting *x* = *I*(*t*), *y* = *S*(*t*), *P* (*x*) = *β/NI*(*t*) + *α ≡ ξ*(*t*) and *Q*(*x*) = 0 *I*(*t*):

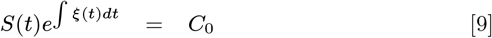

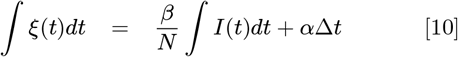

where *α, β, N* are the SEIQRDP model parameters in Eq.1, *C*_0_ is a constant and Δ*t* is the integration range. For the entire time series from start *t* = 0 to a current point *t* = *τ*, Eq.10 is simplified to Eq.11:

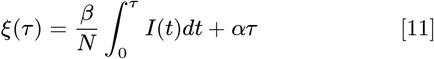

and for *τ* = 0 the *C*_0_ constant is defined as *S*(0) *e ∫*^*ξ*(0)*dt*^ = *S*(0) = *C*_0_. According to the SEIQRDP formulation in Figure 13, the *ατ* (independent of *I*(*t*)) term means that asymptotically all ‘susceptible’ *S*(*t*) will be transferred to ‘insusceptible’ *P* (*t*) compartment regardless of the convergence of ‘infections’ *I*(*t*) asymptotically to zero. This is not realistic for fixed-sized populations, since the infection will probably fade out before everyone gets immunized (or dead), i.e., with some still remaining in *S*(*t*), due to the ‘herd immunity’ phenomenon (1, 2).

Each of the parameters of SEIQRDP model in the block diagram of Figure 13 and the corresponding system of differential equations provide very important hints regarding the dynamics of the underlying system, i.e., the evolution of the epidemic that the model describes, given enough data are available as ground truth. Since the SEIQRDP system of differential equations described by Eq.1 through Eq.7 indicate a specific underlying ‘structure’ for the systems’ dynamics, i.e., solutions to interconnected differential equations, arithmetic Euler-like solution approaches can be used very early on, when the epidemic data are just beginning to accumulate on a day-to-day basis. This is perhaps the main advantage over other approaches that require a statistically significant pool of epidemic data as ground truth from the beginning, e.g., in order to estimate posterior probabilities, etc.

The application of a SEIQRDP model for Greece is presented in section 5.D, while evaluation of a similar model for the entire region is presented in section 6.D.

## 5. Outbreak status in Greece

In order to analyze the progress of the outbreak in Greece, a detailed dataset of daily reports must be used. More specifically, the base dataset used in this study is the one provided for open-access^†^ and updated on a daily bases by John Hopkins CSSE (19), which is the most popular and reliable source at the moment. It includes confirmed cases of Infected *I*(*t*), Deaths *D*(*t*) and Recovered *R*(*t*), registered per-country and in some cases per-province/region/state, collected by the official sources from WHO, CDC (US), European CDC, state authorities in each country, as well as other open-access sources.

The basic curves for Greece from the beginning of the national epidemic are presented in Figures 14 and 15, in linear and logarithmic scale, respectively. From the basic curves, especially the *I*(*t*), it is clear that the progress of the outbreak in Greece is exponential, as expected, but with a relatively slow rate. In the first two weeks there are several step-wise ‘pauses’ and ‘bursts’, as it is usually observed in the early stages of a fast-growing epidemic. This is due to the fact that the set sizes are still relatively small and statistics still unstable, as well as the lack of strict mitigation measures that are usually activated with some delay.

**Fig. 14.**
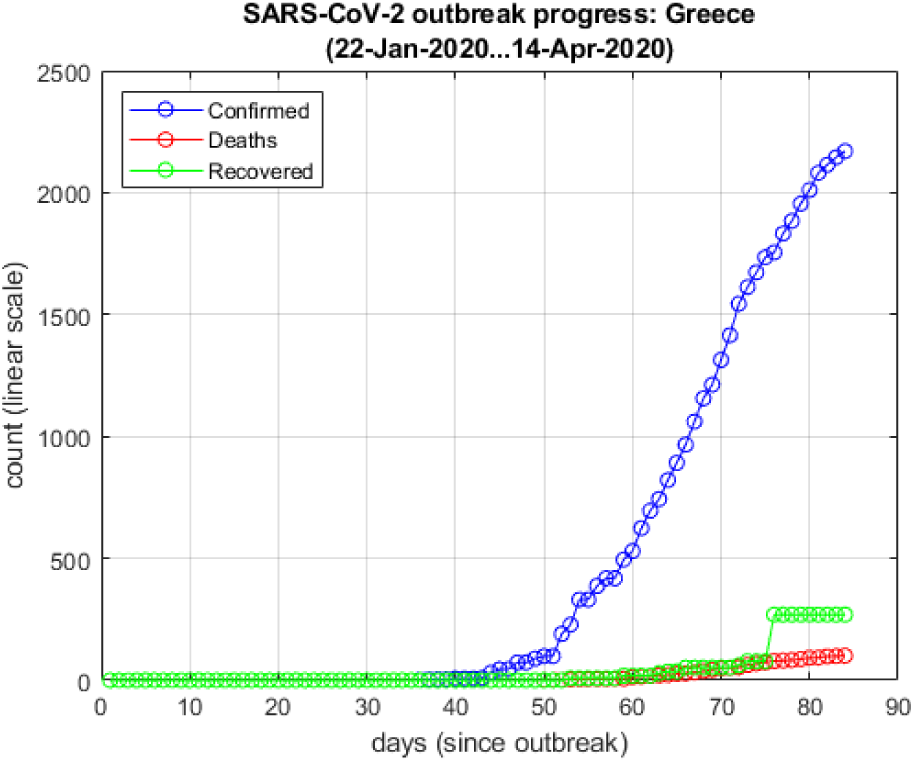
Greece: Infected *I*(*t*), Deaths *D*(*t*) and Recovered *R*(*t*), in linear scale.

**Fig. 15.**
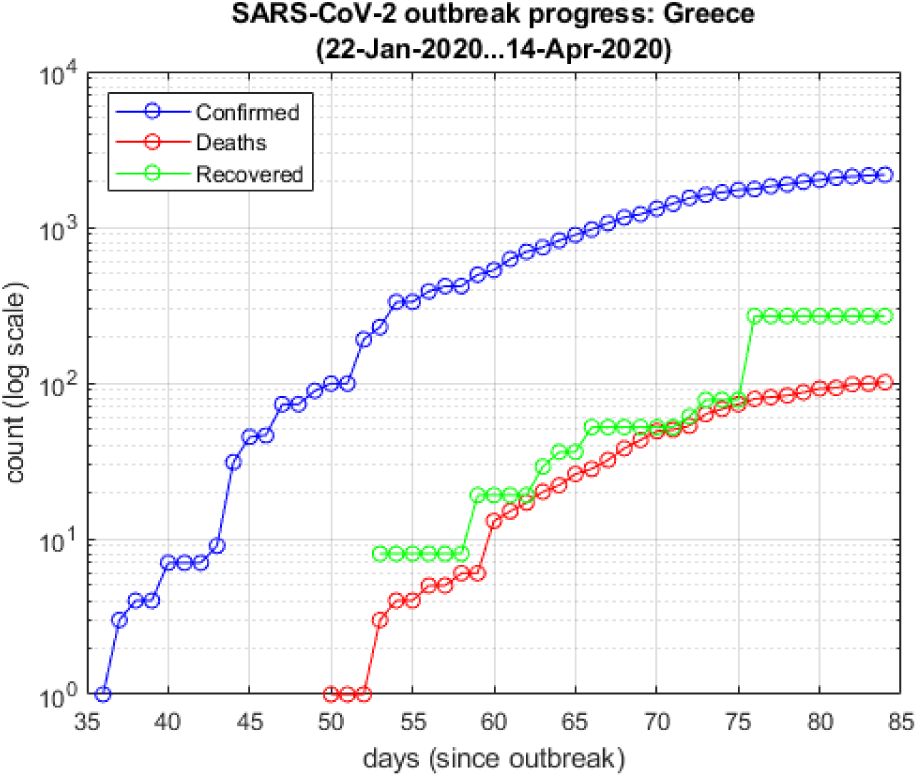
Greece: Infected *I*(*t*), Deaths *D*(*t*) and Recovered *R*(*t*), in logarithmic scale.

The critical ‘necks’ before large increases in new infections seem to be around day 43 and day 51 since the start of the time series, i.e., March 4th and March 12th, respectively. As presented previously in section A, these dates coincide with two important events in the Greek timeline: (a) the 1st confirmed case in the group of tourists who returned from Israel & Egypt and (b) 10 new confirmed cases of unknown origin and the 1st virus-associated death. For (a) it was understood that this group of tourists were asymptomatic for a few days and, thus, have been spreading the virus in the general population in the meantime, as the data proved subsequently. For (b) it was clear that the new confirmed cases of unknown origin, i.e., unrelated to both the group of tourists from Israel & Egypt as well as the other group that returned from northern Italy a few days earlier, were to increase rapidly, as was indeed the case. This is particularly important, because at this point it is understood that the virus has escaped the strict and detailed backtracing of infections and their close encounters, in order to impose targeted isolations. Thus, the operational plan should change to wide-range mitigation measures, as quickly as possible. Fortunately, this is what happened in the following days and, given that the outbreak continued to spread at a fast rate, Greece went into a nation-wide lockdown 11 days later.

### A. Comparison to other countries

Taking into account the progress of the pandemic world-wide since early January 2020, the curves in Figures 14 and 15 reveal that Greece had a slow start. However, it is not sufficient to simply align the offset of each country’s specific event to the others’, e.g., the first reported death or the 100th confirmed case of COVID-19. The entire curve has to be compared and ‘aligned’ to others for a more realistic match. For this reason, Dynamic Time Warping (20, 21) was used for comparing Greece’s infections *I*(*t*) curve to countries in the general region of Europe and central/eastern Mediterranean Sea, especially those in direct ‘contact’ with Greece via international flights.

Figure 16 illustrates the original and the DTW-matched curves of Greece (blue) versus Switzerland (red). The DTW matching provides two important parameters for further study: (a) a more realistic estimate of the time offset for temporal alignment and (b) a DTW-based Euclidean distance that can be used as a similarity measure. Using these two parameters, Figure 17 illustrates the relative temporal difference (offset) and Figure 18 the DTW-based similarity of Greece-versus-others regarding the confirmed cases of infections *I*(*t*).

**Fig. 16.**
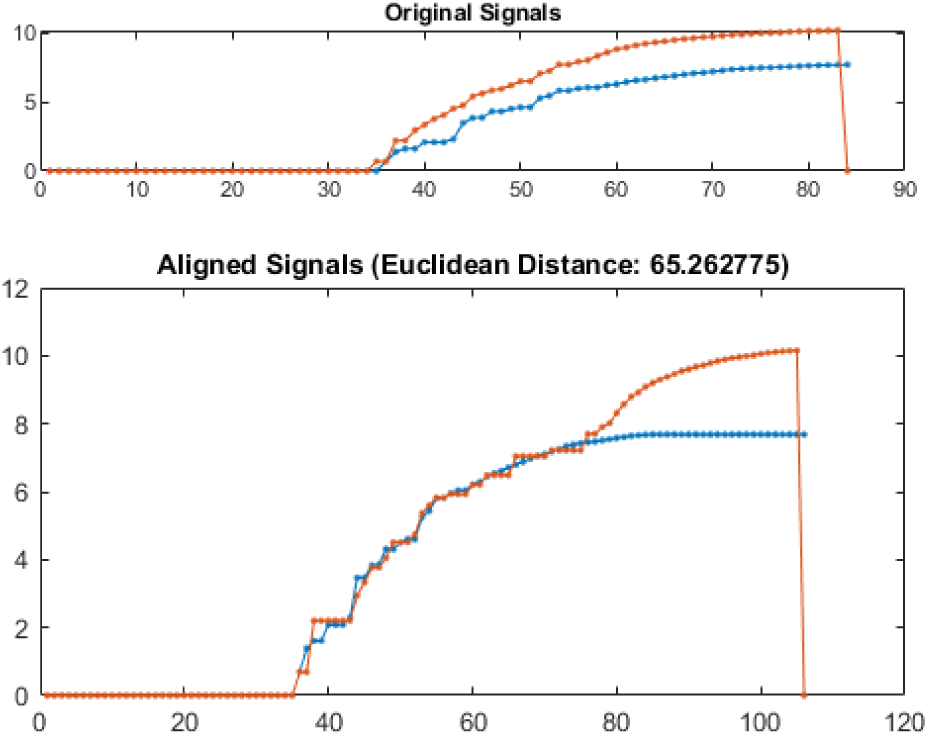
Example of DTW transformation with Euclidean distance of the confirmed infections *I*(*t*) in Greece (blue) versus Switzerland (red).

**Fig. 17.**
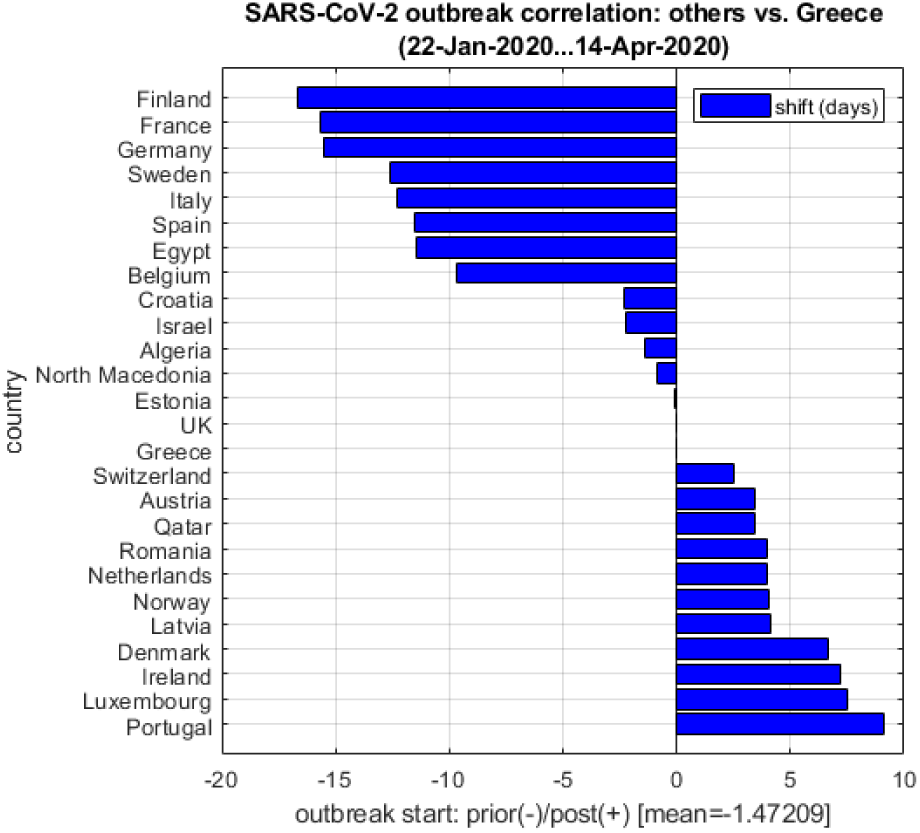
DTW-based temporal offsets of Greece-versus-others regarding the confirmed infections *I*(*t*).

**Fig. 18.**
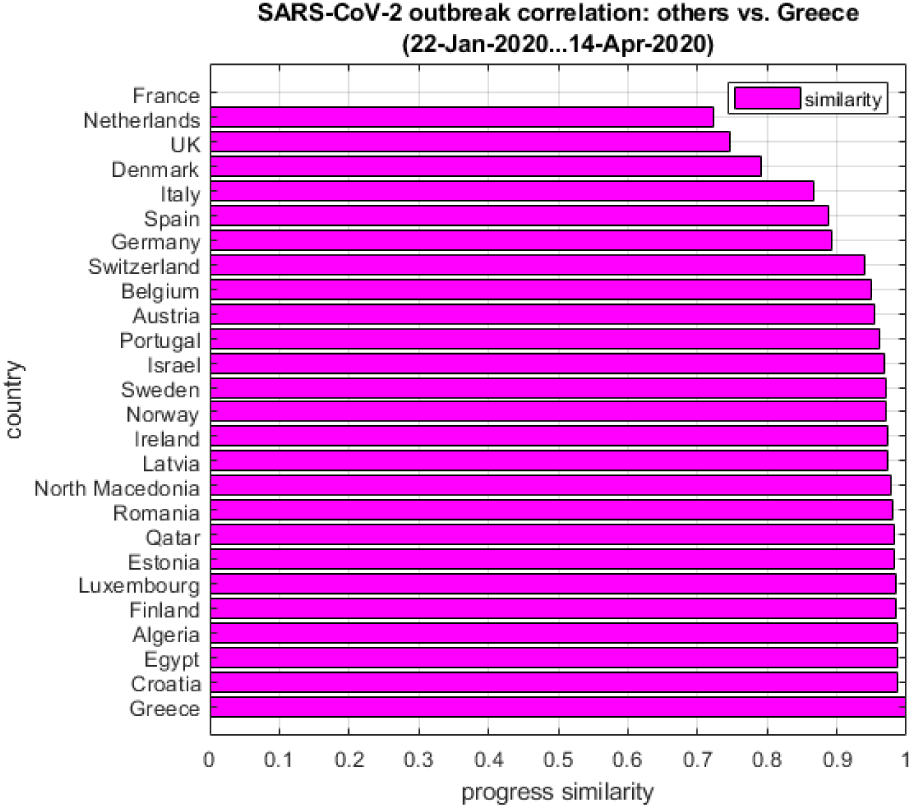
DTW-based similarity of Greece-versus-others regarding the confirmed infections *I*(*t*); similarity here is defined as the max-normalized [0…1] Euclidean distance w.r.t. the entire group.

As the comparative plots in Figures 17 and 18 show, the outbreak in Greece seems more ‘aligned’ with UK, Estonia, and North Macedonia and more ‘similar’ to Croatia, Egypt, Algeria, Finland, etc. It should be noted that at the beginning of the outbreak in Greece the temporal lag offset w.r.t. the group of countries presented here was more than 14 days ‘behind’. During the last two weeks it seems that the outbreak in Greece gets gradually ‘synchronized’ with what is happening with the rest of the countries in this region and now the lag seems to be less than two days (*t* = −1.47). This is particularly important in terms of a similar alignment of the timeline of gradual deactivation of mitigation measures: If a country is still far ‘behind’ in the progress of the epidemic, then re-establishing international flights increases the risk of having another wave of the outbreak inside this country, as discussed earlier in section 3.

### B. Data-driven analytics

Using the daily time series for confirmed cases of Infected *I*(*t*), Deaths *D*(*t*) and Recovered *R*(*t*) in Greece, appropriate approximations can be developed in order to estimate important parameters of the outbreak in Greece.

Specifically, using the *I*(*t*) data series the following exponential formulation can be designed according to Eq.12:

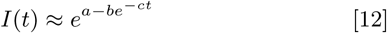

where *a, b, c* are the function parameters. Their best-fit optimal values in the least-squares (LSE) sense (22) and the 95% confidence intervals for Greece are presented in Table 1.

**Table 1.** LSE-optimal function parameters in Eq.12 for Greece.

| Parameter | optim.value | conf.interval |
| --- | --- | --- |
| $a$ | 8.097 | (7.974, 8.221) |
| $b$ | 8.722 | (8.420, 9.024) |
| $c$ | 0.064 | (0.060, 0.068) |

Note: $b$ and $c$ are used with a negative sign in Eq.12. All confidence intervals are calculated for $p = 0.95$ . Goodness-of-fit: $R^2=0.992$ , RMSE=0.017

Figure 19 presents the difference between a ‘naive’ exponential *y*(*t*) = *e*^*cx*(*t*)^ (red) with only one parameter and the good fit (*R*^2^=0.992, RMSE=0.017) of Eq.12 (magenta). In the second case it is clear that the function design, i.e., an exponentially saturating formula, as well as a linear error weighting that is applied towards the more recent date range, provides the very robust set of optimized parameters that Table 1 presents. It should also be noted that, as the *I*(*t*) becomes more linear due to the gradual slow-down of the national outbreak, the exponential factor *c* in Eq.12 and Table 1 converges to zero and factors *a* and *b* increase, i.e., the value of *I*(*t*) asymptotically approaches its upper threshold as it crossed well inside phase 5a of the epidemic and moves towards its ‘peak’ (see: section 3).

**Fig. 19.**
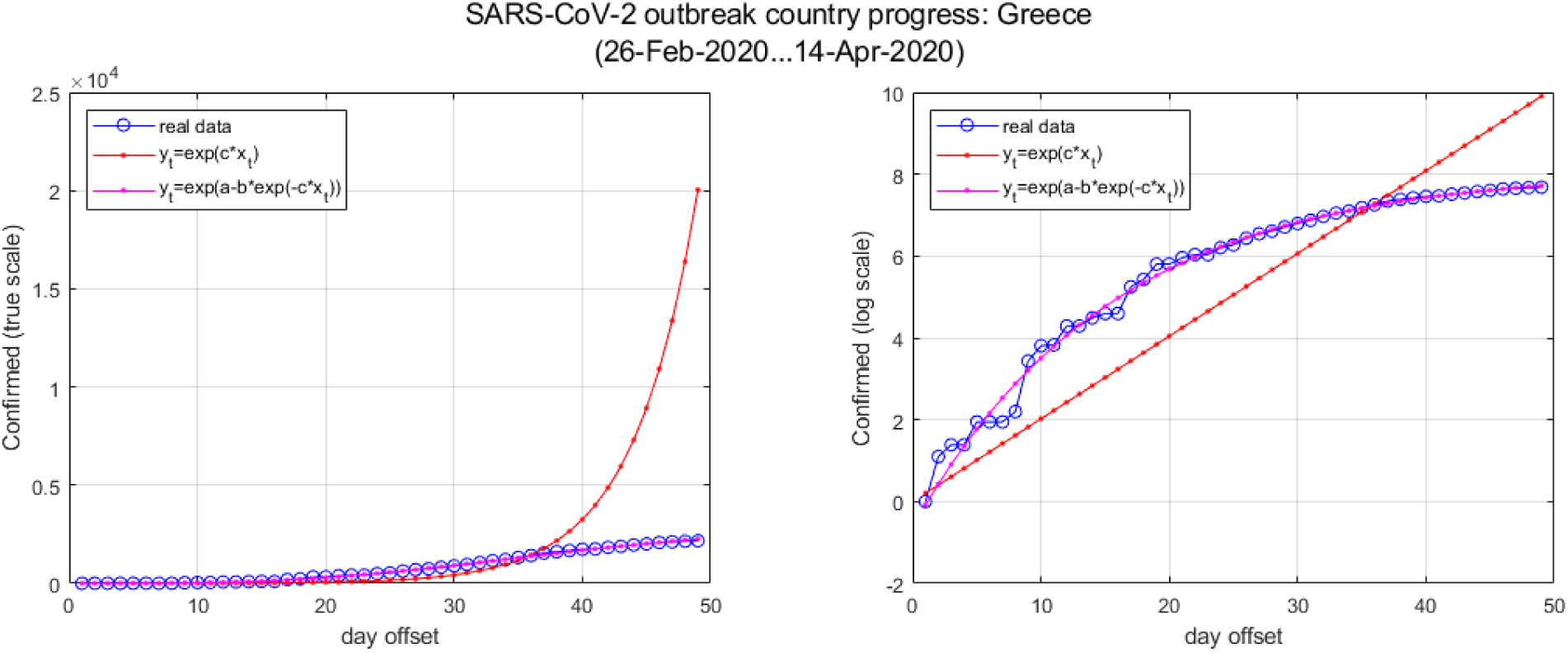
‘Naive’ exponential *y*(*t*) = *e*^*cx*(*t*)^ (red) with only one parameter and the good fit of Eq.12 (magenta) for Greece. (*R*^2^=0.992, RMSE=0.017)

It should be noted that the approximation of Eq.12 is valid only for epidemic phases 4 and most of phase 5a as explained in section 3, because after the curvature sign change or ‘inflection point’^‡^ (23, 24) of *I*(*t*) at the start of phase 5 (*d*^2^*I*(*t*)*/dt*^2^ = 0) and especially when closing towards the peak between sub-phases 5a and 5b, the curvature *d*^2^*I*(*t*)*/dt*^2^ becomes negative and the slope *dI*(*t*)*/dt* of *I*(*t*) begins to decrease, i.e., the saturation point of Eq.12 is no longer an asymptotic limit as it ‘leaves behind’ the actual *I*(*t*) peak.

Using the approximation of Eq.12 and Table 1, the *basic reproduction number R*_0_ can be approximated by an analytical formulation too, taking the limit of the mean ratio of the marginal increase of *I*(*t*), as described in Eq.13. Figure 20 illustrates the progress of the (estimated) *R*_0_ for Greece from the beginning of the national epidemic, according to Eq.13.

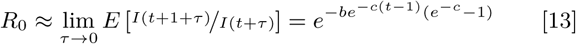

**Fig. 20.**
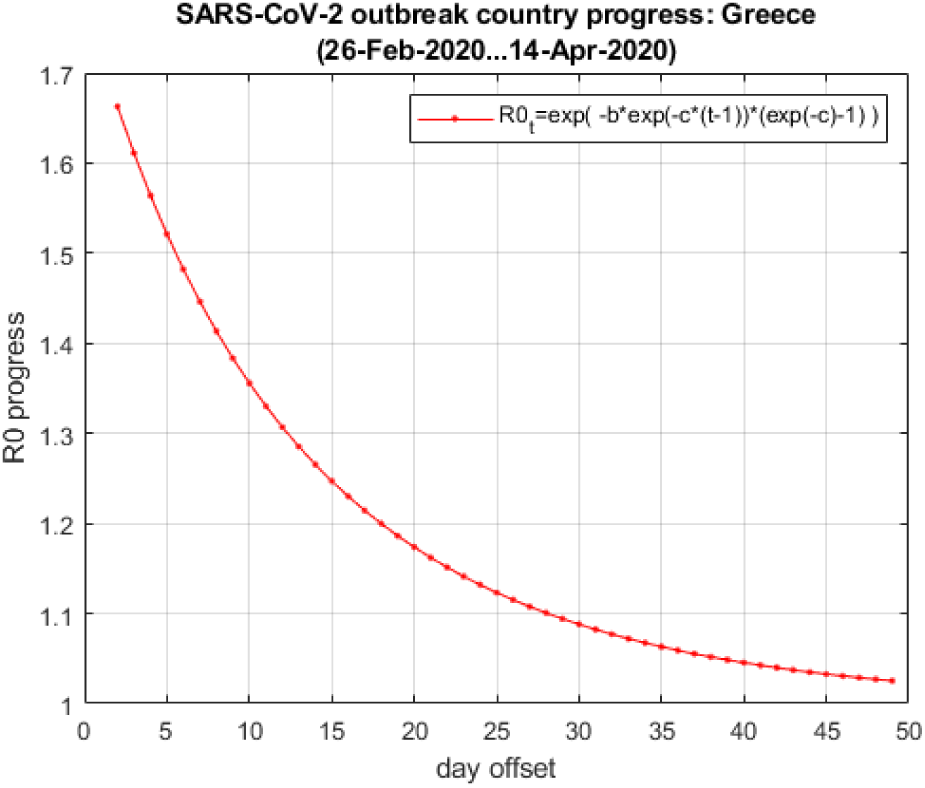
Progress of the (estimated) *R*_0_ and *Dd* for Greece from the beginning of the national epidemic, according to Eq.13.

The estimate of *R*_0_ in Figure 20 at the current state (far right) can be confirmed by taking a sliding-window linear regression (LR) (22, 25) on the most recent temporal window on the actual *I*(*t*) data series, using the standard LR formulation as described in Eq.14, Eq.15 and Eq.16. Additionally, the corresponding doubling rate *Dd* can be estimated from *R*_0_ by Eq.17.

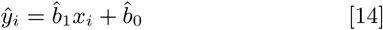

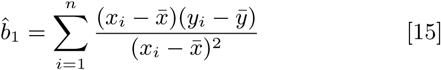

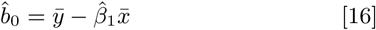

where 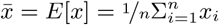 and 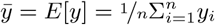.

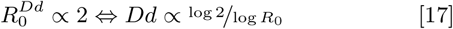

According to the *R*_0_ approximation of Eq.13 and using a marginal step of *τ* = 1 day, the current eight-day LR approximations of *R*_0_ = 1.03 (1.03, 1.04) and *Dd* = 22 (18, 25) in Figure 21 confirm the corresponding estimations calculated by Eq.13 and Eq.17. The value of *R*_0_ seems to have reached the critical low threshold of 1.00 briefly on 11-12th of April, followed by consecutive days of stable value of 1.03 until today. It is worth noting that the Greece’s team of experts on the official daily briefing of April 14th stated for the first time that *‘R*_0_ *is now falling below 1’*.

**Fig. 21.**
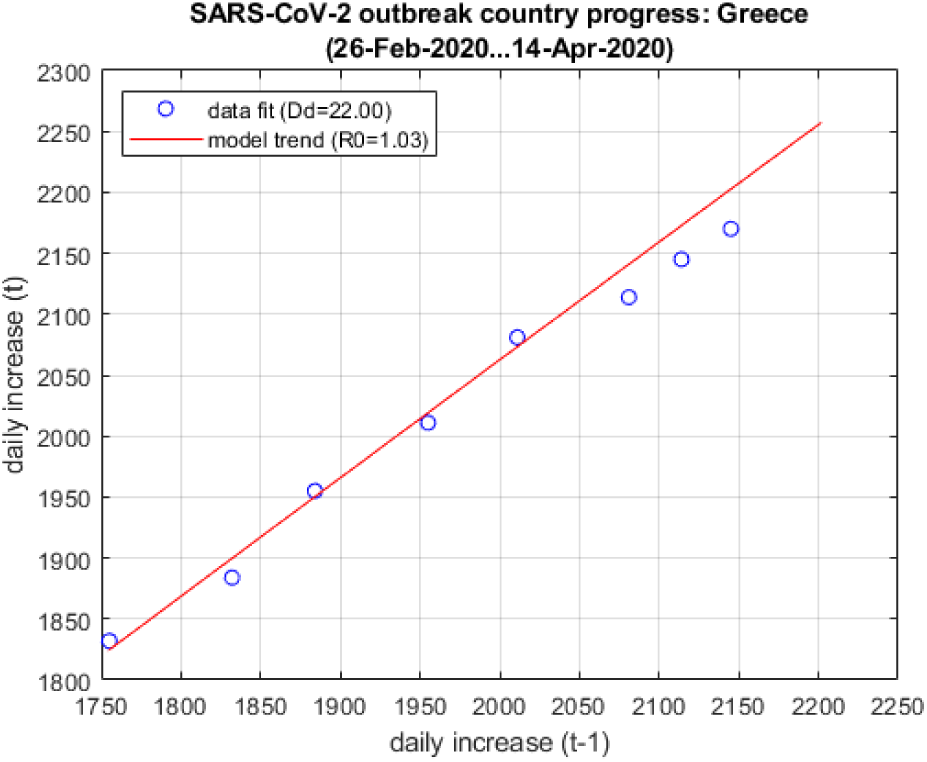
Progress of the (estimated) *R*_0_ and *Dd* for Greece, according to the current eight-day LR approximations for *R*_0_ and Eq.17 for *Dd*.

Two other important parameters that can be estimated by taking a sliding-window LR on the most recent temporal frame on the actual *I*(*t*) data series, using the formulation as described in Eq.14, Eq.15 and Eq.16, are the ratio of ICU to *I*(*t*) and the ratio of Deaths *D*(*t*) to *I*(*t*). The first is a good indicator of whether there is a trend towards possible saturation of the health system in treating severe cases of infection. The second can be used to compare the national fatality rate with the global statistics estimated internationally, in order to see if the under-reporting of *I*(*t*) is much more significant than in other countries and, hence, verify the inherent risk of all the infections-related assumptions in the target country.

Fortunately, the ICU trend in Greece seems to be steadily decreasing during the last ten days, after a sharp rise the days before. Given the fact that Greece entered this crisis with only 550 and still with less than 990 ICU currently available throughout the country, keeping this number down is crucial in the quality of treatment of severe cases and of course avoiding the saturation of the health system if it comes close to its maximum capacity. As Figure 22 shows, the current estimation of the ICU-to-*I*(*t*) ratio has dropped from 4-5% and is now at about 3.5% (76 in 2170) and with steady 10-day LR slope (−0.044).

**Fig. 22.**
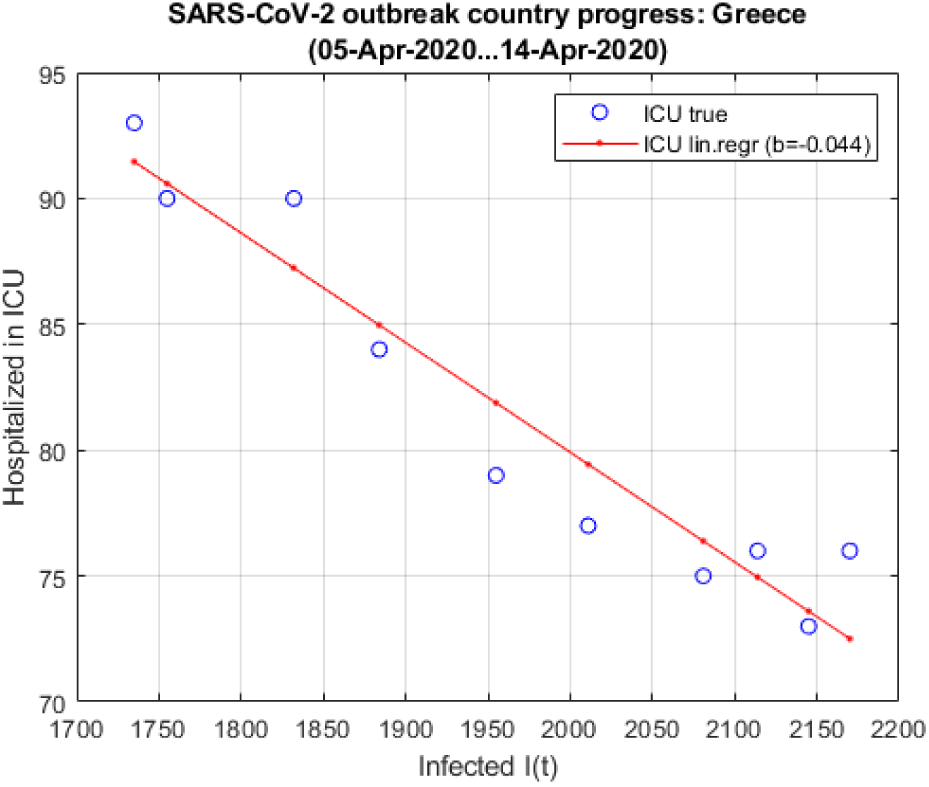
Progress of the (estimated) ICU rate for Greece, according to the current 16-day LR approximation against *I*(*t*).

The current *D*(*t*)*/I*(*t*) ratio for Greece is also providing a good LR fit, presented in Figure 23, with a value fluctuating slightly between 2% and 6%, currently at 4.65% (101 in 2170) and with relatively steady 4-day LR slope (+0.085). This is generally inside the ‘global’ region of 2% to 5% that is estimated internationally for COVID-19, as presented previously in Figure 8.

**Fig. 23.**
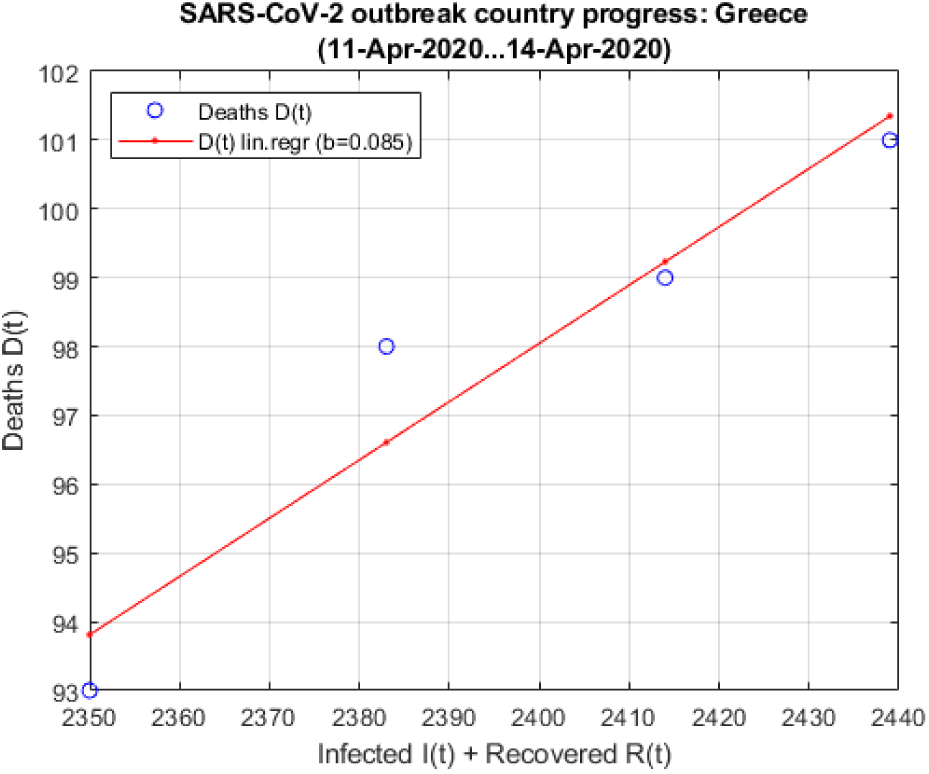
Progress of the (estimated) fatality rate for Greece, according to the current 4-day LR approximation against *I*(*t*).

It has been accepted by state officials that the underreporting of *I*(*t*) in Greece may be up to 20:1 (only 1 in 20 infections registered) or more, given the numbers from other countries and the targeted-only tests in Greece. A recent study^§^ (26) estimates the true number of *I*(*t*) for Greece between 9,652 and 20,377 in the general population^¶^ rather than 2,145 officially documented by April 13th, i.e., an underreporting rate of 6.5:1 in currently active infections. Nevertheless, the *D*(*t*)*/I*(*t*) fatality ratio is still within the expected zone, indicating that the overall tracking of *I*(*t*), *D*(*t*) and *R*(*t*) data series are reliable enough to track the general progress of the national outbreak, at least as much as in any other country still behind or just entering phase 5 of its national epidemic.

## C. Modelling the epidemic

For a more in-depth analysis of the basic data curves *I*(*t*), *D*(*t*) and *R*(*t*) of the COVID-19 epidemic in Greece and in any other country, the corresponding time series have to be investigated in terms of linear and periodic trends, i.e., analyze them into their primary frequency components. In signal processing this is normally done via time-domain filtering or a Fourier transformation for frequency-domain filtering (27). In case of non-stationary signals, filters can be formulated via adaptive algorithms in order to track the stationarity shifts of the underlying event (28).

A more generic alternative that is usually applied in complex systems where the input/output association is governed by multiple ‘internal’ parameters is via state-space representation, where the input and output are connected through an internal ‘state vector’. In this case, the model is trained so that for specific inputs produces specific outputs based on functional dependencies between each of them and the state vector. If the training data are saturated with noise or if the system itself is non-stationary, this formulation can be generalized in a probabilistic way - this is the core idea behind Kalman filtering (28).

In almost all cases of analytical formulations for signal processing the basic assumption is that the underlying system is a linear model or can be closely approximated within a useful range by ‘linearization’ (28). Unfortunately, these methods are not easily applicable per-se in long-term epidemic modelling, since the underlying system in governed by a system of differential equations that describe the dynamics of the phenomenon, which in epidemic outbreaks is the non-trivial interactions between groups of populations including the susceptible, the exposed, the infected, the fatalities, the recovered, etc. More on this approach is presented later on in section 4.

One important factor that can be investigated regardless of the long-term approximation of the underlying system is analyzing the step-wise dependencies between successive data points, especially the confirmed cases *I*(*t*). In statistical terms, this is done by estimating the auto-correlation in the time series for various lags, which produces a quantitative description of dependencies between dates, i.e., separated by a specific number of days. A more descriptive analysis is the Fourier transformation (FFT) (27) of the daily *I*(*t*) increase, i.e., the time series of new confirmed cases of infections, revealing the spectral components of the spread of the virus in the population.

Figure 24 presents the normalized auto-correlation plot of Δ*I*(*t*) (blue); it is evident that the two sides of the lobe are almost entirely linear and the LR lines (red) in each side are almost parallel to the reference line (green) that corresponds to the case of Δ*I*(*t*) = *c*, i.e., for constant daily increase of *I*(*t* + *τ*) = *cτ* + *I*(*t*). This proves that, at least in asymptotic behaviour towards the current state, *I*(*t*) increase in Greece is gradually becoming linear.

**Fig. 24.**
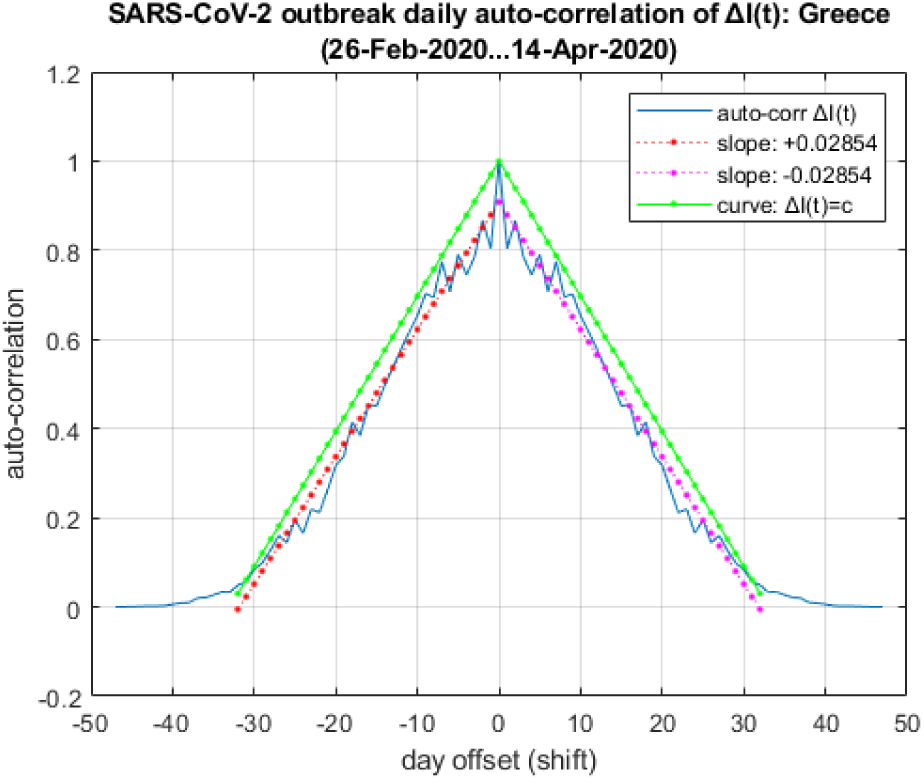
Normalized auto-correlation plots of Δ*I*(*t*) (blue), the LR lines (red) in each side and the reference line (green) of the case of Δ*I*(*t*) = *c* (constant).

Additionally, using FFT for spectral decomposition, Figure 25 presents the spectrum (normalized, abs.value, half-width) of Δ*I*(*t*) (blue). From left to right, the x-axis corresponds to temporal window size of the corresponding component. In practice, this means that the left-most points in the plot correspond to spectral components spanning to a temporal window of half the entire time series, i.e., periodic trends of three weeks or more. On the opposite side, the right-most points in the plot correspond to spectral components spanning to a temporal window of 1, 2, 3 days, etc. From this plot it is clear that there are indeed two groups of significant spectral components: (a) short-term in the span of 4 and 6 days and (b) long-term in the span of 20 days. This is observation coincides with two very important periodic trends of the COVID-19 outbreak, already documented empirically throughout the world (29, 30): (a) the incubation period of the virus, mostly asymptomatic and thus highly contagious, is estimated at 5.1-5.2 days; and (b) the suggested quarantine safe period for the onset of symptoms in asymptomatic carriers or after their recovery is at least two weeks, i.e., associated with the long-term spectral component if incubation period is included (infection just before quarantine ends). Observation (a) can probably be associated with the short-term spectral component of days 4-6, i.e., the window between the onset of an infection and the manifestation of disease symptoms. Observation (b) may be associated with the long-term spectral component around day 20, given that the additional week can be attributed to infections set right at the end of a two-week quarantine.

**Fig. 25.**
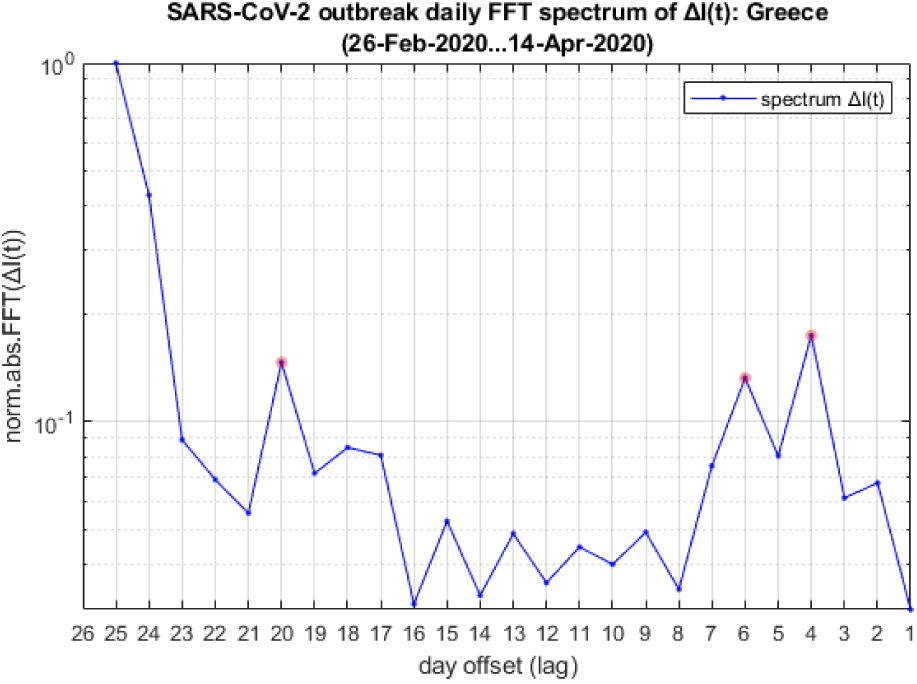
FFT spectrum (normalized, abs.value, hald-width) of Δ*I*(*t*) (blue) in logarithmic scale; marked data points (red) are the primary spectral components.

It should be noted that spectral components describe both increasing and decreasing trends, i.e., either sudden increases *and* decreases of Δ*I*(*t*). Thus, first observation suggests that large changes of Δ*I*(*t*) w.r.t the mean are expected with a frequency of about the 4-6th day, as well as the 20th day. Data are still limited for a long-term (statistically significant) validation per-component, but observed fluctuations of these primary spectral components are limited and, hence, these numbers can indicate valid hints regarding the effectiveness of specific mitigation measures activated during that time.

Another way to track the periodic ‘bursts’ of newly reported infections as the are reported on a daily basis is to track the changes in the short-term slope of Δ*I*(*t*). Instead of approximating the entire *I*(*t*) curve as in Eq.12 and Figure 19 for estimating the long-term behaviour, a short-term temporal window can be used to approximate the LR slope of *I*(*t*) as it progresses, i.e., the amplitude and sign of Δ*I*(*t*) changes over few subsequent days. Figure 26 illustrates such a short-term tracking of Δ*I*(*t*) via the 1st-order differential 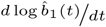 of LR slope of *I*(*t*) with 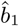 as defined in Eq.15, or in other words the Δ^2^*I*(*t*), over a short-term sliding window of four days.

**Fig. 26.**
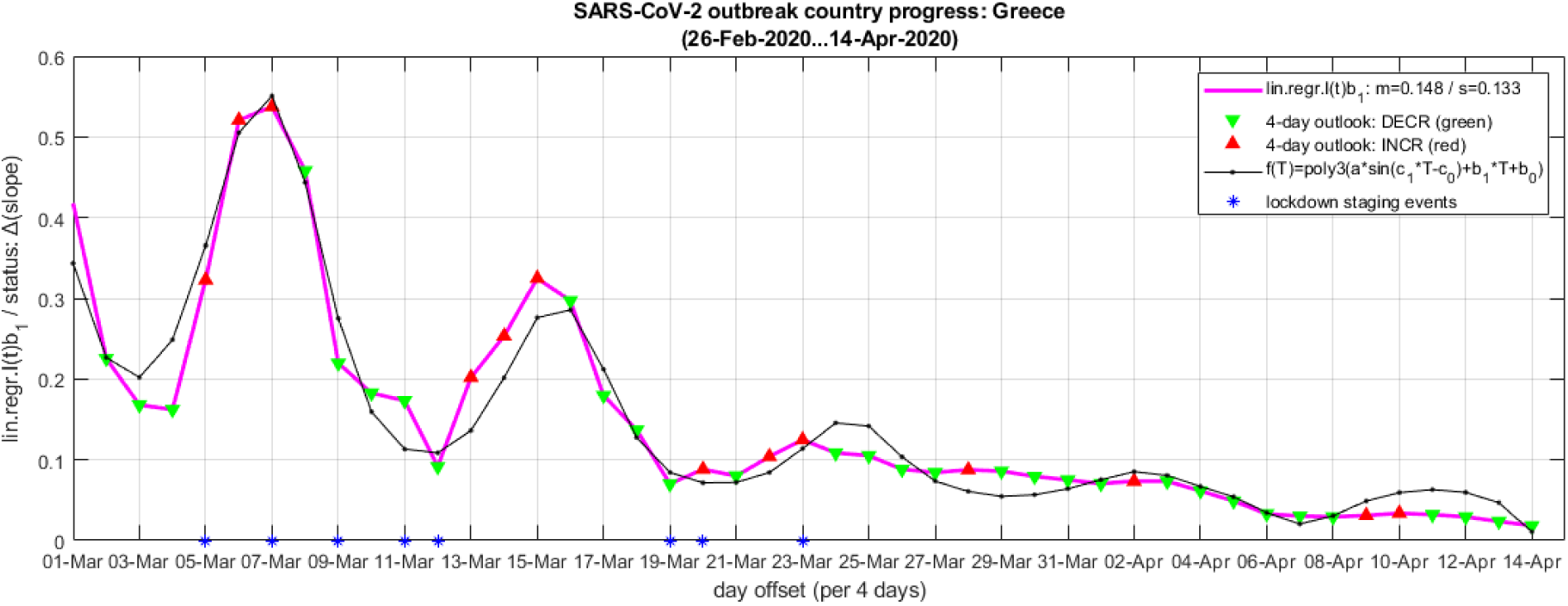
4-day sliding window slope 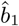 Δ*I*(*t*) (magenta), annotations of increasing/decreasing daily trends (red/green) and LSE-fitted approximation of the 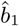.

The main curve (magenta) in Figure 26 is the short-term LR slope 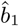 value for *I*(*t*) as it evolves; the arrow annotations indicate decreasing (green) or increasing (red) trends; the asterisks (blue) on the x-axis indicate the major events regarding the activation of mitigation measures in Greece as described in the timeline in section A. Finally, the asymptotically fading sinusoid (black) is a LSE-fitted approximation of *g*_(_*t*) by *ĝ* (*t*), as defined by Eq.18, Eq.19 and Eq.20, with their LSE-optimal parameters presented in Table 2.

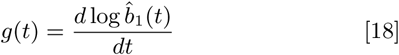

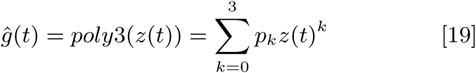

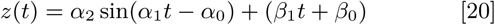

**Table 2.** LSE-optimal function parameters in Eq.19 and Eq.20 for Greece.

| Parameter | optim.value | conf.interval |
| --- | --- | --- |
| $\alpha_2$ | 0.070 | (0.040, 0.100) |
| $\alpha_1$ | 0.713 | (0.681, 0.745) |
| $\alpha_0$ | 0.003 | (-0.978, 0.985) |
| $\beta_1$ | -0.008 | (-0.020, 0.004) |
| $\beta_0$ | 0.359 | (0.128, 0.590) |
| $p_3$ | 19.330 | (11.220, 27.440) |
| $p_2$ | -3.575 | (-7.223, 0.074) |
| $p_1$ | 0.430 | (0.001, 0.859) |
| $p_0$ | 0.044 | (0.025, 0.063) |

Note: $\alpha_0$ is used with a negative sign in Eq.20. All confidence intervals are calculated for $p = 0.95$ .

From the approximation curve in Figure 26 there is clear indication of three major factors: (a) periodic trend, captured by the first part of Eq.20 with *α*_*i*_ parameters, (b) linear decreasing trend, captured by the second part of Eq.20 with *β*_*j*_ parameters, and (c) asymptotically fading trend, captured by the 3rd-degree polynomial of Eq.19 with *p*_*k*_ parameters. The periodic trend parameters, more specifically the *α*_1_ = 0.713, can be translated from radians to daily temporal range via *α*_1_*/*2*π* = *t/T* where *T* = 49*d* is the current length of the data series for Greece (April 14th) since the first confirmed infection case (February 26th), hence yielding a period *t*_*p*_ = ^*α*^_1_^*T*^*/*2*π≈* 0.011348*T≈* 5.56*d*. Again, this number almost coincides with the empirical data regarding the incubation (asymptomatic) period of COVID-19, estimated at 5.1-5.2 days (29, 30). Additionally, the linear trend with marginally downward slope (*β*_1_ *<* 0) shows a gradual slowdown of the ‘force’ of the outbreak in Greece, while the asymptotically fading trend captured by the 3rd-degree polynomial (*p*_*k*_) shows that the ‘bursts’ or ‘waves’ of the newly reported infections (Δ*I*(*t*)) are also fading out after less than ^*T*^*/t*_*p*_ *≈* 8.81 periods (5-6 in practice, according to Figure 26). Taking into consideration that the main mitigation measures (blue asterisks in x-axis) in Greece were activated just before each major ‘peak’ in (Δ*I*(*t*)), it can be stated that they were imposed in a timely manner and, thus, they were appropriate and effective in containing the ‘force’ of the national outbreak until today.

### D. SEIQRDP model for Greece

Based on the compartmentalized SIR/SEIR epidemic modelling described in section 3, in this study a SEIQRDP model was designed and trained using the *I*(*t*), *D*(*t*) and *R*(*t*) data series for Greece, from 26-Feb-2020 to 14-Apr-2020. The purpose of this work was to re-estimate the general properties of SARS-CoV-2 virus and the COVID-19 outbreak on the national level, in order to: (a) confirm that the available data series are adequate and their evolution in accordance to the research outcomes from other countries and on world-wide level; and (b) to estimate the progress of the outbreak on the mid-/long-term level, specifically for the epidemic phases, the expected peak date and magnitude, etc.

The solution of the SEIQRDP system of differential equations defined in Eq.1 through Eq.7 was estimated by a standard LSE solver (22) for iterative matching of the predicted trajectories to the real data. For inclusion of a minimal under-reporting of infections, *I*(*t*) data series was amplified by an additional +8% upon its recorded values. The LSE solver used centralized differential estimators, 1e-6 time step size (86.4 ms) and 1e-6 error tolerance for stopping criterion. In all cases, the process converged to a solution within less than 30 iterations, which hints that the actual epidemic data for Greece are well-described by the SEIQRDP model.

Figures 27 and 28 present the best-fit solution of the SEIQRDP model (points) and its projection (lines) until August (2020), in linear and logarithmic scale, respectively. The dotted line in Figure 28 illustrates the onset of a subsequent surge of the outbreak if all the mitigation measures (quarantine) was to be deactivated immediately (April 15th). Table 3 presents the values for all the SEIQRDP parameters for the best-fit solution.

**Table 3.** LSE-optimal SEIQRDP model parameters in Eq.1 through Eq.7 for Greece (26-Feb-2020 to 14-Apr-2020), minimal underreporting of infections.

| Parameter | optim.value |
| --- | --- |
| $\alpha$ | 0.1030 |
| $\beta$ | 3.0000 |
| $\gamma$ | 0.1186 |
| $\delta$ | 0.0352 |
| $\lambda$ | 2.9986 |
| $\kappa$ | 0.0468 |

Note: $I(t)$ amplified for 1.08:1 marginal under-reporting; LSE used centralized differential estimators, 1e-6 time step size (86.4 ms), 1e-6 error tolerance for stopping criterion; convergence in <30 iterations; starting conditions: $E_0=0$ , $I_0=1$ , $D_0=0$ , $R_0=0$ ; estimated population (2018): $N_{pop}=10,724,599$ .

**Fig. 27.**
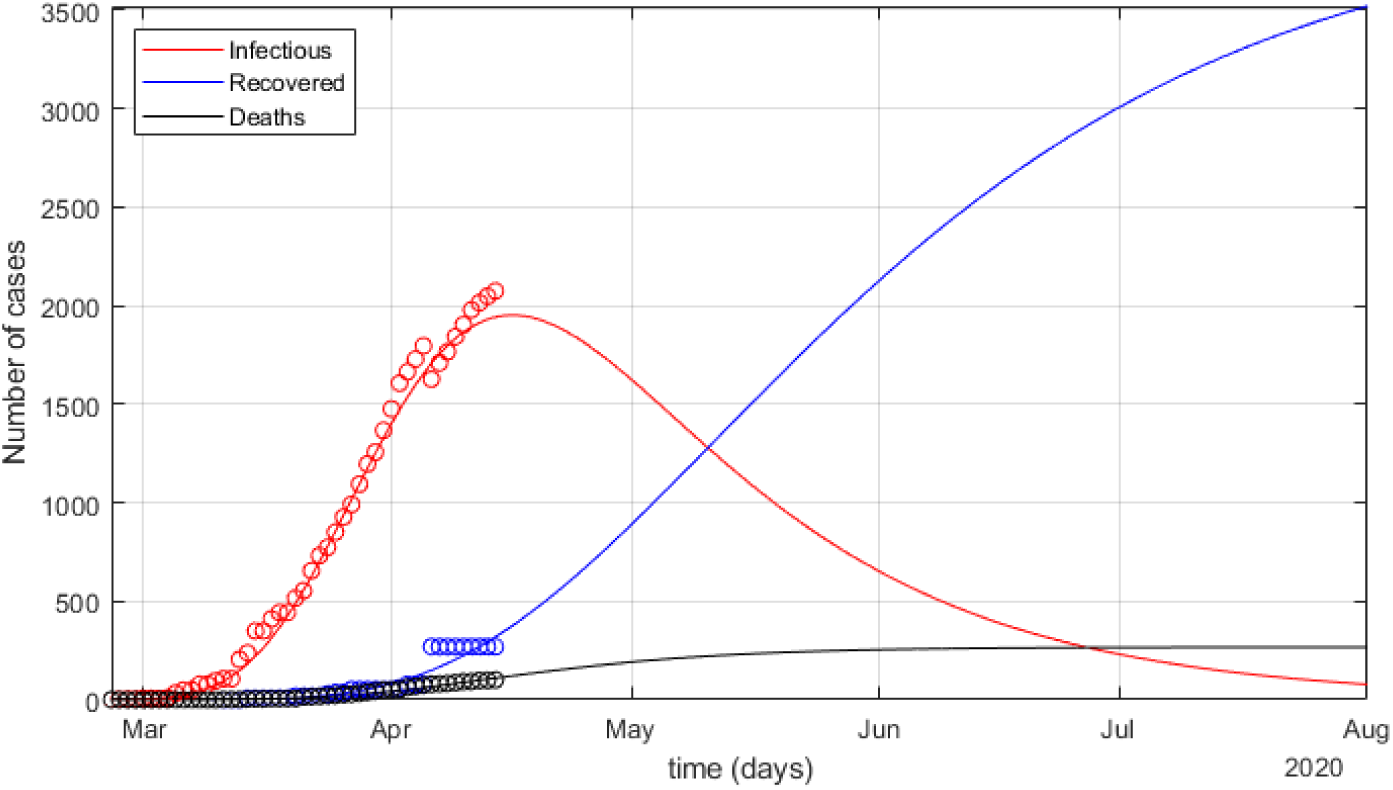
SEIQRDP best-fit model (points) and its projection (lines) for marginally (1.08:1) under-reported *I*(*t*), *D*(*t*) and *R*(*t*) until August (2020) in linear scale.

**Fig. 28.**
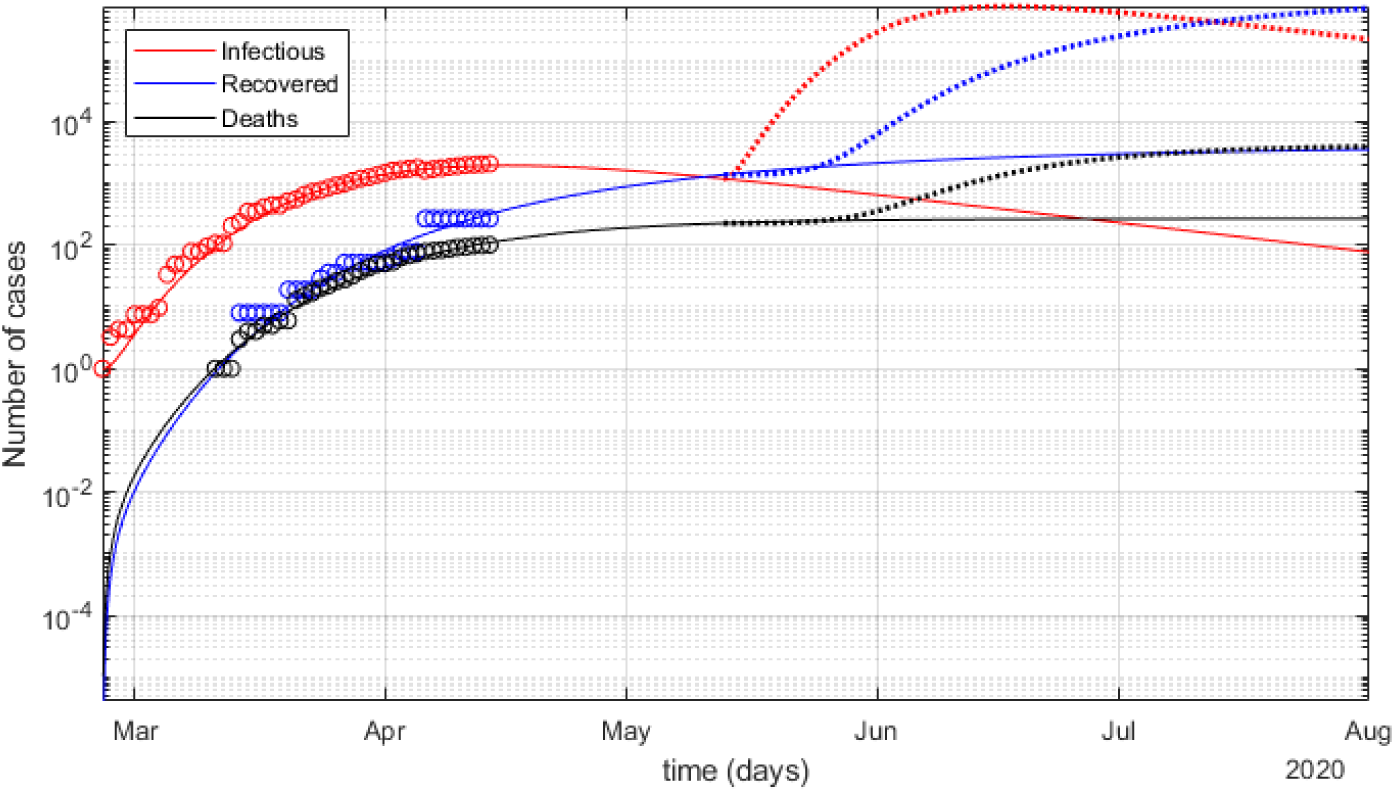
SEIQRDP best-fit model (points) and its projection (lines) for marginally (1.08:1) under-reported *I*(*t*), *D*(*t*) and *R*(*t*) until August (2020) in logarithmic scale.

Given the estimated under-reporting of confirmed cases *I*(*t*) as explained earlier in section 5.B, two additional scenarios were tested: one for the low estimation of 4.5:1 (9,652/2,145) and one for the high estimation of 9.5:1 (20,377/2,145).

Similarly to the previous, Figures 29 and 30 present the best-fit solution of the SEIQRDP model (points) and its projection (lines) until August (2020), in linear and logarithmic scale, respectively, for the ‘low’ scenario of 4.5:1 under-reporting of *I*(*t*). The dotted line in Figure 30 illustrates the onset of a subsequent surge of the outbreak if all the mitigation measures (quarantine) was to be deactivated immediately (April 15th). Table 4 presents the corresponding values for all the SEIQRDP parameters for the best-fit solution.

**Table 4.** LSE-optimal SEIQRDP model parameters in Eq.1 through Eq.7 for Greece (26-Feb-2020 to 14-Apr-2020), ‘low’ under-reporting of infections.

| Parameter | optim.value |
| --- | --- |
| $\alpha$ | 0.1156 |
| $\beta$ | 3.0000 |
| $\gamma$ | 0.1917 |
| $\delta$ | 0.0067 |
| $\lambda$ | **0.5296** |
| $\kappa$ | 0.0380 |

Note: $I(t)$ amplified for 4.5:1 'low' under-reporting; LSE used centralized differential estimators, 1e-6 time step size (86.4 ms), 1e-6 error tolerance for stopping criterion; convergence in <30 iterations; starting conditions: $E_0=0$ , $I_0=1$ , $D_0=0$ , $R_0=0$ ; estimated population (2018): $N_{pop}=10,724,599$ .

**Fig. 29.**
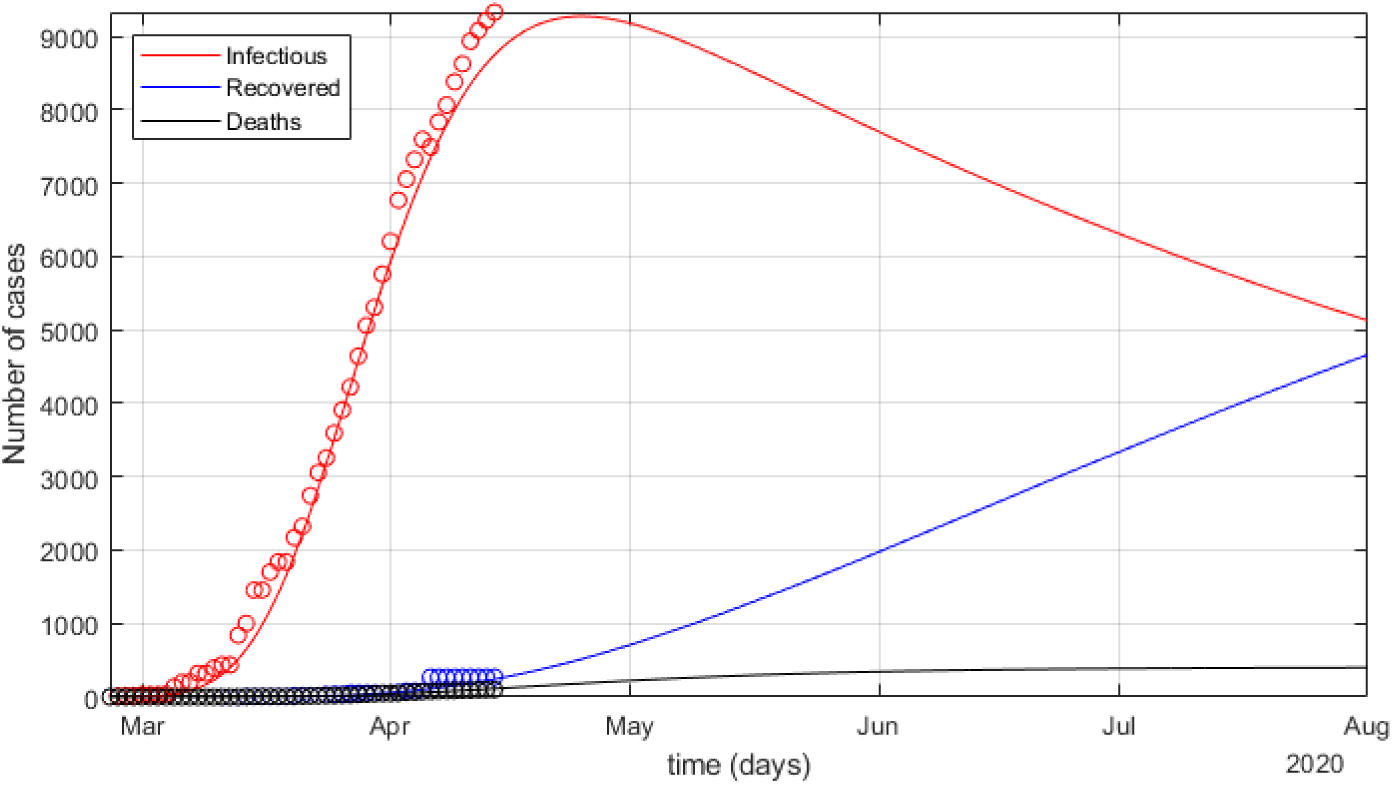
SEIQRDP best-fit model (points) and its projection (lines) for ‘low’ (4.5:1) under-reported *I*(*t*), *D*(*t*) and *R*(*t*) until August (2020) in linear scale.

**Fig. 30.**
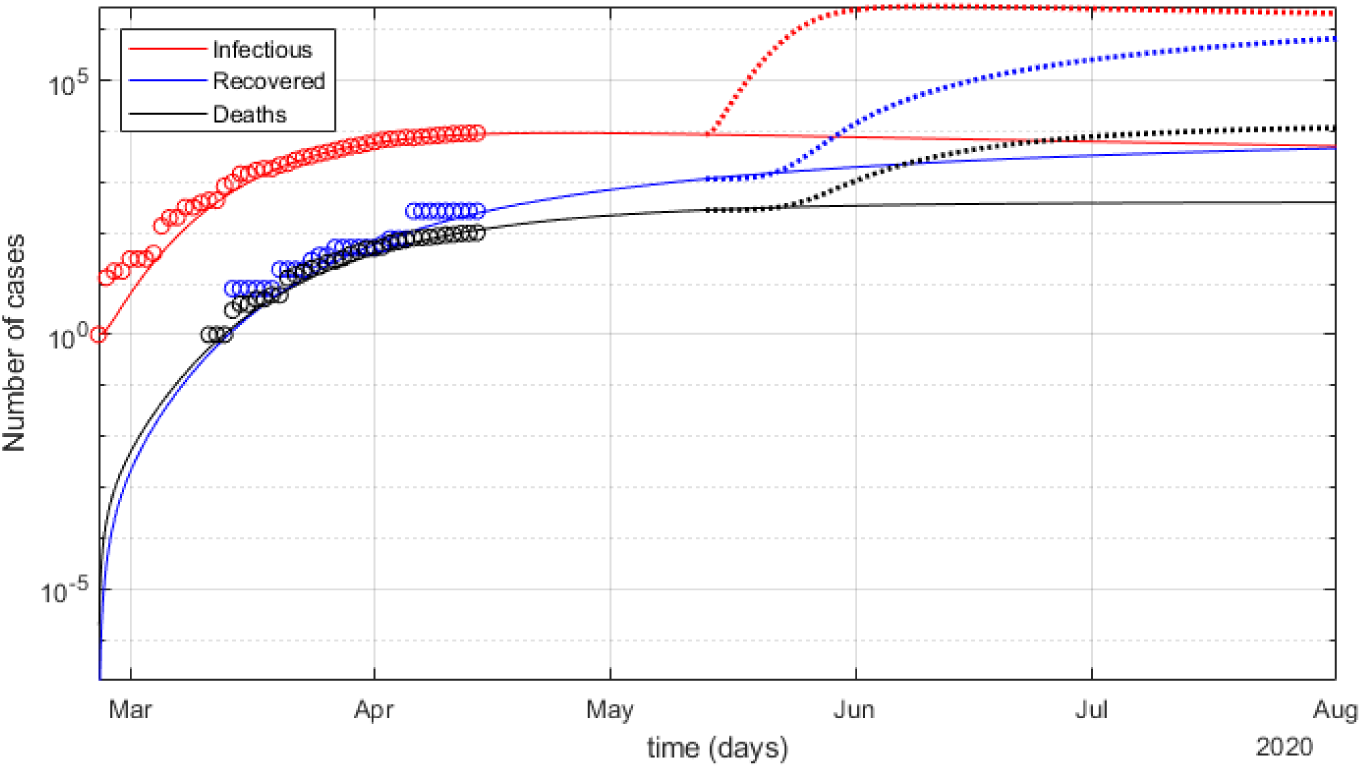
SEIQRDP best-fit model (points) and its projection (lines) for ‘low’ (4.5:1) under-reported *I*(*t*), *D*(*t*) and *R*(*t*) until August (2020) in logarithmic scale.

Finally, Figures 31 and 32 present the best-fit solution of the SEIQRDP model (points) and its projection (lines) until August (2020), in linear and logarithmic scale, respectively, for the ‘high’ scenario of 9.5:1 under-reporting of *I*(*t*). The dotted line in Figure 32 illustrates the onset of a subsequent surge of the outbreak if all the mitigation measures (quarantine) was to be deactivated immediately (April 15th). Table 5 presents the corresponding values for all the SEIQRDP parameters for the best-fit solution.

**Table 5.** LSE-optimal SEIQRDP model parameters in Eq.1 through Eq.7 for Greece (26-Feb-2020 to 14-Apr-2020), ‘high’ under-reporting of infections.

| Parameter | optim.value |
| --- | --- |
| $\alpha$ | 0.1180 |
| $\beta$ | 3.0000 |
| $\gamma$ | 0.2435 |
| $\delta$ | 0.0013 |
| $\lambda$ | **3.0000** |
| $\kappa$ | 0.0241 |

Note: $I(t)$ amplified for 9.5:1 'high' under-reporting; LSE used centralized differential estimators, 1e-6 time step size (86.4 ms), 1e-6 error tolerance for stopping criterion; convergence in <30 iterations; starting conditions: $E_0=0$ , $I_0=1$ , $D_0=0$ , $R_0=0$ ; estimated population (2018): $N_{pop}=10,724,599$ .

**Fig. 31.**
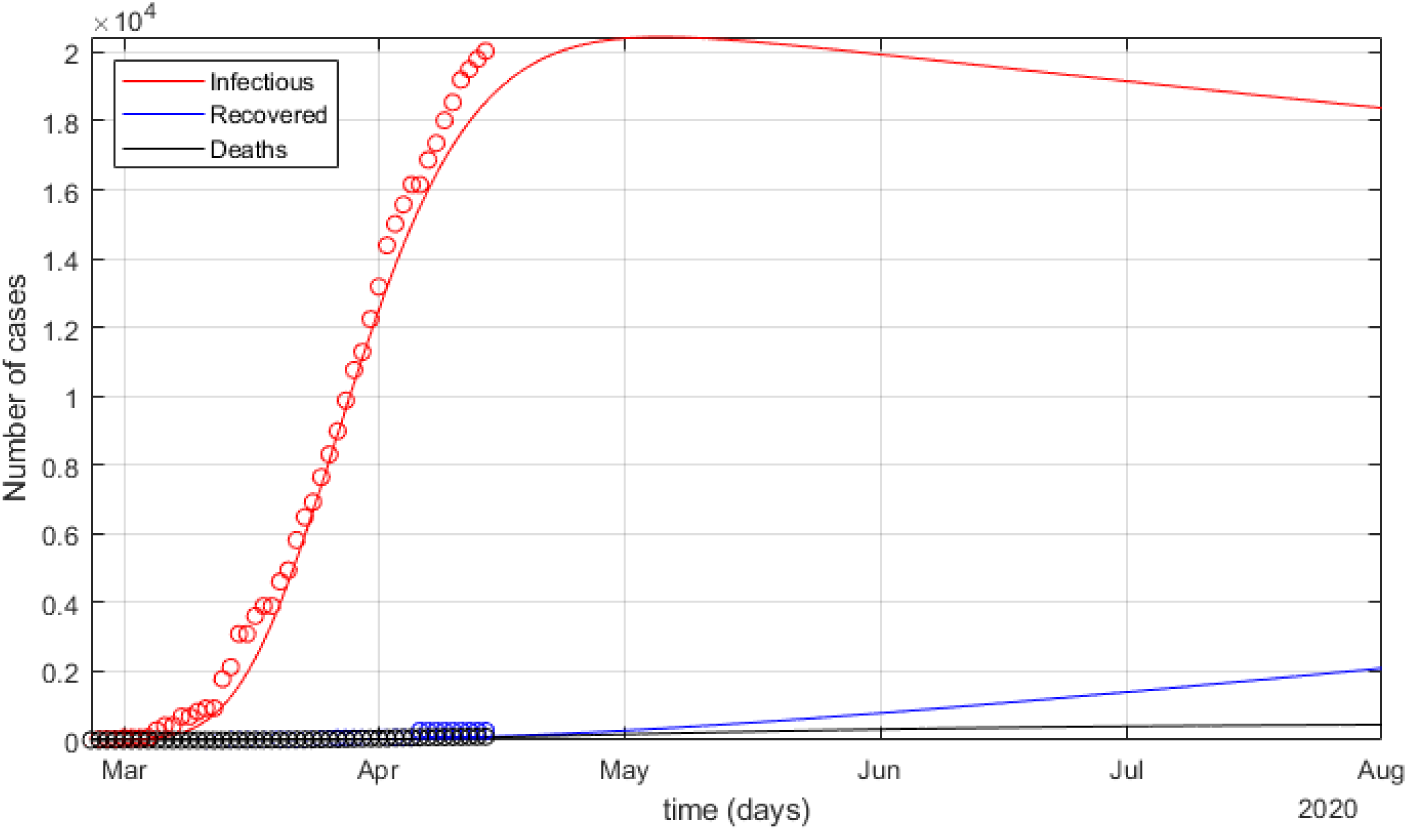
SEIQRDP best-fit model (points) and its projection (lines) for ‘high’ (9.5:1) under-reported *I*(*t*), *D*(*t*) and *R*(*t*) until August (2020) in linear scale.

**Fig. 32.**
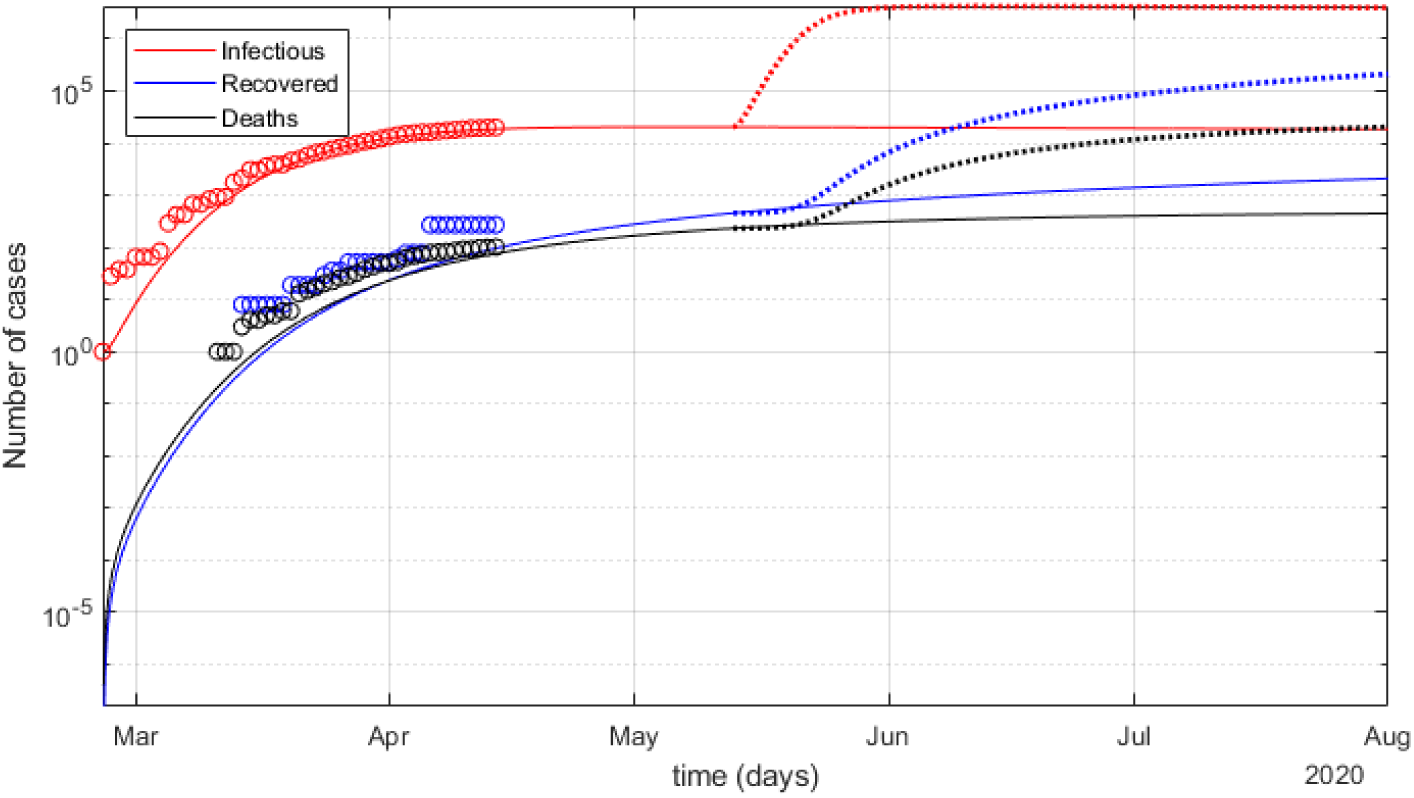
SEIQRDP best-fit model (points) and its projection (lines) for ‘high’ (9.5:1) under-reported *I*(*t*), *D*(*t*) and *R*(*t*) until August (2020) in logarithmic scale.

In Tables 4 and 5, parameter *λ* is highlighted as the only one with significant variation between the three under-reporting scenarios for *I*(*t*). More specifically, the marginal 1.08:1 and the ‘high’ 9.5:1 solutions are compatible, but the ‘low’ 4.5:1 is not, since its *λ* value is almost six-fold smaller than the other two, despite the fact that in all cases the LSE best-fit solutions are of good quality (see comments). The *lambda* parameter in SEQRDP is associated to the rate of transfers between ‘quarantined’ *Q*(*t*) and ‘recovered’ *R*(*t*) compartments (see: Figure 13). This is an interesting issue for further investigation at a later time, when more data will be available through post-analysis after the resolution of the national outbreak, in relation to a more realistic estimation of the actual under-reporting of *I*(*t*) during the crisis.

Summarizing the SEIQRDP best-fit solutions for the various scenarios of infections under-reporting, estimations of peak *I*(*t*) dates for Greece are presented in Table 6. Note that, given the epidemic phase uncertainty due to the still limited data series, no value estimations are presented for *I*(*t*). Nevertheless, the goodness-of-fit of the SEIQRDP solutions provides a valid ‘explanation’ for the dynamics of the national epidemic, i.e., the interaction between the compartments. Thus, the overall shape and scale of the corresponding curves can be considered as safe for general assessments, including peak *I*(*t*) dates.

**Table 6.** SEIQRDP projections for Greece based on the currently available epidemic data (26-Feb-2020 to 14-Apr-2020), for various scenarios of infection under-reporting.

| Under-reporting scenario for $I(t)$ | Peak $I(t)$ date |
| --- | --- |
| none (1.0:1) | 15-Apr |
| marginal (1.08:1) | 16-Apr |
| 'low' (4.5:1) | 25-Apr |
| 'high' (9.5:1) | 5-May |

Note: Given the epidemic phase uncertainty due to the still limited data series, no value estimations are presented for $I(t)$ .

## 6. Discussion

The data analytics, best-fit model parameters and projected outcomes for Greece, as presented in the previous sections, provide solid evidence that the COVID-19 outbreak at the national level can be tracked with adequate accuracy for the general assessment of the situation, including the transition through the phases of the epidemic. Additionally, it can be compared to the corresponding world-wide analytics and epidemic parameters for validation.

### A. Outlook for Greece

Officially, the SARS-CoV-2 spread in Greece begun with the first confirmed case of infection on February 26th. From there on, both the infections are rising monotonically, i.e., the national epidemic is still prior to its peak, where the infections are expected to be stabilized. Additionally, the rate of increase in the confirmed infections has been rising too, thus yielding an exponential growth, up to a point somewhere between days 71-73 past January 22nd, i.e., somewhere in April 2nd-4th (see: Figure 14. Taking into account the evolution of the outbreak and the progress of the estimated epidemic parameters beyond this point, it seems that the rate of increase has been gradually slowing down until today. In fact, there have been three consecutive days in offset 81+ past January 22nd of decreasing slope in the rate of confirmed infections (April 12th-14th: *I*(80 + *τ*) = {+33, +31, +25}, *τ* = {1, 2, 3}). This is evidence that, indeed, a rising inflection point was passed around April 2nd-4th and, the rate of infections increase is slowing down and Greece is nearing its peak. In other words, it is now beyond phase 4 and well inside phase 5a, according to the descriptions in section 3.

Based on this evidence and the fact that ICU needs (recently at 70 − 80 and steady) are still well inside the currently available capacity of the country’s health system (*≈* 990), ICU availability can be safely considered as guaranteed with a very high probability through the outbreak peak and beyond that. In the long-term, the SEIQRDP projection estimates the total number of fatalities directly related to COVID-19 at a few hundreds at most up to and including the entire summer, provided that the national epidemic will be tracked continuously and accurately, as well as an effective plan for adapting the mitigation measures appropriately in the next several months. On the opposite side, high-impact mitigation measures and prompt containment of an epidemic most commonly translates to a low immunization / high residual susceptibility level for the general population after the outbreak, provided that no large-scale artificial immunization (vaccination) will be available soon and re-introduction of the virus in Greece is almost a certainty after the international travelling restrictions are gradually deactivated. Hence, additionally one or more outbreak surges are expected in the national level within the year, always in correlation to what is happening to neighbouring countries and international travelling, as well as the speed of deactivation of national mitigation measures (lift of quarantine). In the end, a high proportion of the general population in Greece has to get immunized through recovery or vaccination, in order to establish an evolutionary stable ‘herd immunity’ state.

An additional negative factor is the expected slow fadeout period, i.e., longer phase 6 in the epidemic, due to the estimated high rate of under-reported infections. Results from the corresponding SEIQRDP best-fit projections of peak dates in Table 6 show that the higher the under-reporting rate, the slower the progress of the outbreak is through its peak and fade-out period. This is a very important aspect of planning the deactivation of mitigation measures at the national level, as well as the need to very strict border checking regarding international travelling and large-scale COVID-19 testing in the general population for accurate tracking of the residual or new spread.

### B. Supporting evidence

In more technical terms, the evidence that leads to the outlook for Greece can be seen in the results presented in previous sections. More specifically, the close inspection of Figures 14 and 15, as well as the details in the plot of ‘status tracking’ in Figure 26, reveal that the national epidemic is indeed slowing down. Additionally, the epidemic parameters estimated from the related data for Greece are within the expected ranges w.r.t. the international experience with COVID-19 since the start of the pandemic. This is also true for the periodic trends of the national outbreak, based on the main spectral components (see: Figure 25).

The overall ‘positioning’ of Greece in terms of the onset, phase difference and infections curve similarity towards the countries in the greater area (Europe, north Africa, near Middle-East), according to Figures 17 and 18, reveal that the national outbreak is a rather average-case rather than an outlier. The values of parameters in the SEIQRDP best-fit solutions seem to behave as expected (except perhaps *λ*) and the value of its most important ‘exponential spread’ parameter is reliably at *β* = 3.0, regardless of the under-reporting ratio in infections. In general, during the last 7-10 days all the data analytics and modelling parameters seem to be ‘smoothing out’ gradually towards a more steady state, i.e., approaching the peak of confirmed infections, thus ending phase 5a and beginning phase 5b (downwards slopes).

### C. Factors of uncertainty

Despite the fact that most of the evidence presented for Greece can be considered reliable, there are still some aspects that require verification or further investigation, perhaps in a post-analysis level after the resolution of the current crisis, when more data will be available.

One such factor of uncertainty is the presence of a significant data shift or ‘step’ in the time series, observed visually on day 76 past January 22nd, i.e., April 6th. This ‘step’ constitutes a very low +20 newly reported infections *I*(*t*) between two days of +62 and +77, while at the same time having a very large daily increase in recoveries *R*(*t*) (see: Figures 14 and 15). This may be attributed to late confirmation or official reporting of clinical tests for patients prior to their release from hospital, but this is only an assumption.

Another uncertainty factor is the actual rate of underreported infections at the national level, mostly due to targeted-only COVID-19 tests instead of wide-range tests in the general population as in other countries. Although several epidemic parameters are not affected by this (e.g., SEIQRDP *b* = 3.0), there are other important aspects that require this clarification (e.g., estimation of peak date), even in terms of post-analysis after the resolution of the current crisis, in order to assess the effectiveness of the policies taken.

Finally, the sub-optimal behaviour of the SEIQRDP modelling w.r.t. the general region around Greece is a prohibiting factor in assessing the ‘next day’ on the international level. Although the national epidemic in Greece is steadily ‘synchronizing’ with that in the rest of the countries, no safe conclusions can be stated for the situation and possible recurrent surges of the outbreak when travelling restrictions will gradually get deactivated.

### D. Outlook for the region

Similarly to the results presented earlier for the national epidemic in Greece, Figures 33 and 34 present the best-fit solution of the SEIQRDP model (points) and its projection (lines) until August (2020), in linear and logarithmic scale, respectively, assuming no under-reporting of *I*(*t*), for the greater region around Greece. The dotted line in Figure 34 illustrates the onset of a subsequent surge of the outbreak if all the mitigation measures (quarantine) was to be deactivated immediately (April 15th). Table 7 presents the corresponding values for all the SEIQRDP parameters for the best-fit solution, which seem to deviate significantly from the corresponding solutions for Greece, except *β ≈* 2.7 which is close to *β* = 3.0 as in Tables 3 through 5.

**Table 7.** LSE-optimal SEIQRDP model parameters in Eq.1 through Eq.7 for the greater region around Greece (24-Jan-2020 to 13-Apr-2020), no under-reporting of infections.

| Parameter | optim.value |
| --- | --- |
| $\alpha$ | 0.0032 |
| $\beta$ | 2.6936 |
| $\gamma$ | 1.9076 |
| $\delta$ | 2.0000 |
| $\lambda$ | 0.0006 |
| $\kappa$ | 0.0787 |

Note: $I(t)$ not amplified for under-reporting; LSE used forward differential estimators, 1e-6 time step size (86.4 ms), 1e-6 error tolerance for stopping criterion; moderate convergence in $\approx 2000$ iterations; starting conditions: $E_0=0$ , $I_0=1$ , $D_0=0$ , $R_0=0$ ; bounding box for region: Lat $\in [19.0, 70.0]N$ , Lon $\in [-11.0, 56.0]E$ ; estimated population (1.e6): $0.9 < N_{pop} < 1.5$ .

**Fig. 33.**
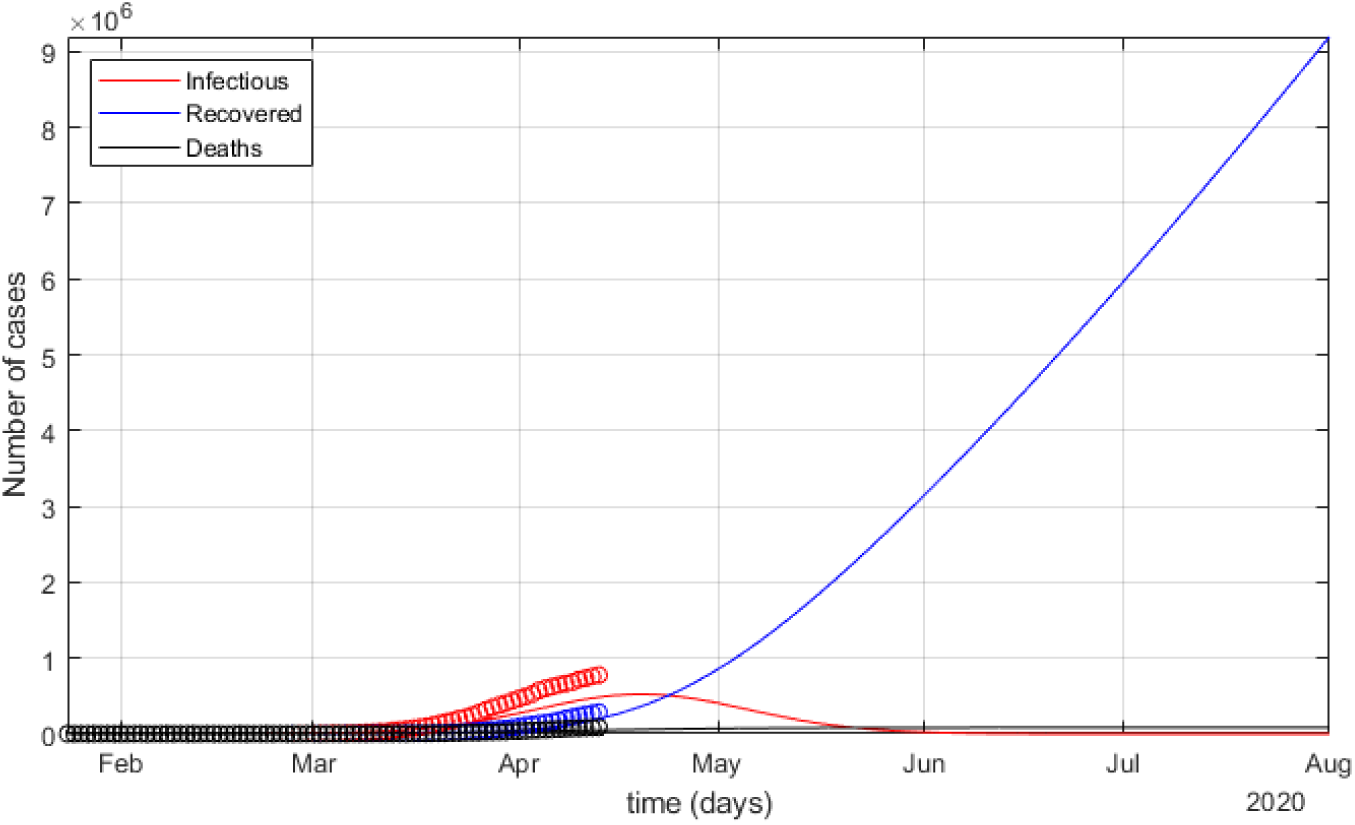
SEIQRDP best-fit model (points) and its projection (lines) for no under-reported *I*(*t*), *D*(*t*) and *R*(*t*) until August (2020) in linear scale, for the greater region around Greece.

**Fig. 34.**
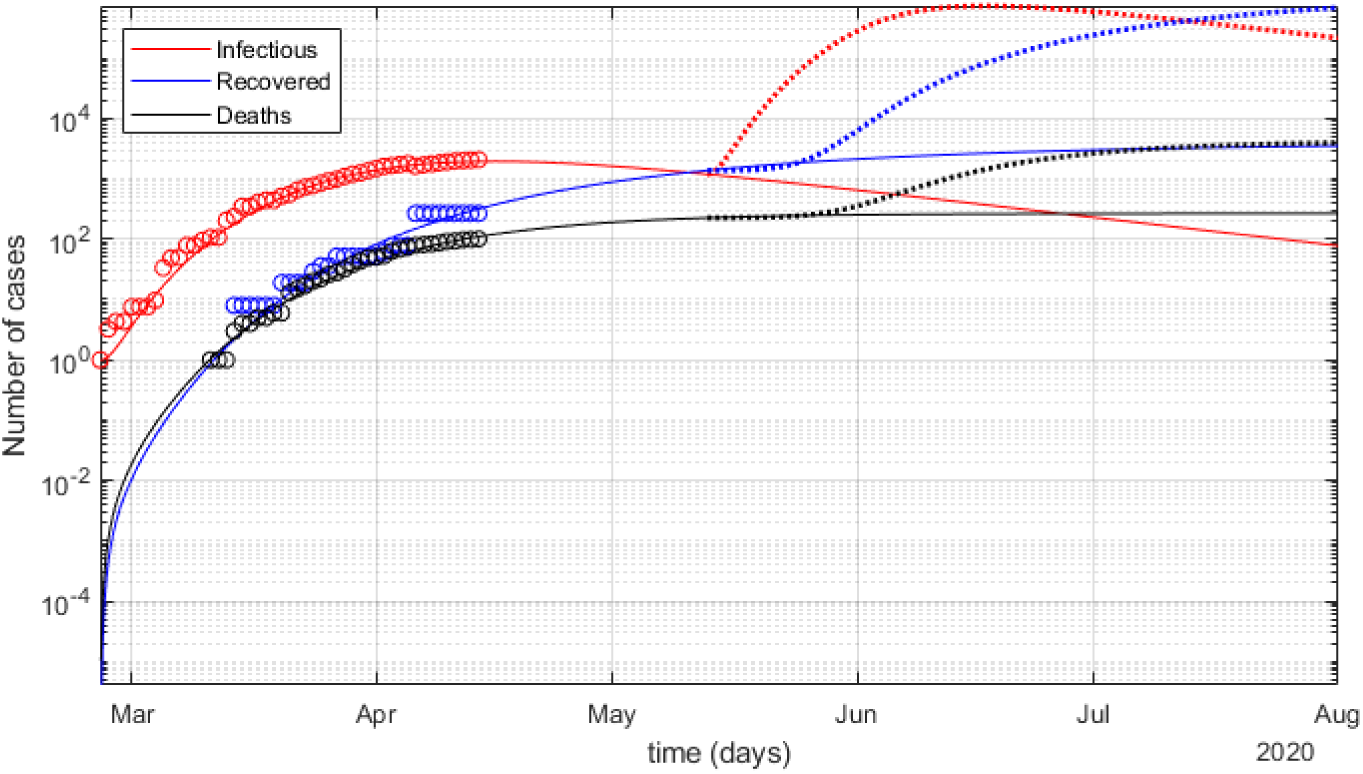
SEIQRDP best-fit model (points) and its projection (lines) for no under-reported *I*(*t*), *D*(*t*) and *R*(*t*) until August (2020) in logarithmic scale, for the greater region around Greece.

As presented previously for Greece, the SEIQRDP best-fit solution for the greater region around Greece provides an estimation of the peak *I*(*t*) date, which seems to be around the 19th of April. Note that, given the epidemic phase uncertainty due to the still limited data series, as well as the moderate-only convergence of the LSE solution process, no reliable value estimation can be provided for *I*(*t*). Nevertheless, Figures 33 and 34 seem to provide a valid ‘explanation’ for the dynamics of the greater-region epidemic, i.e., the interaction between the compartments. Thus, the overall shape and scale of the corresponding curves can be considered as safe for general assessments, including the peak *I*(*t*) date at some point closely after the mid-April. Additionally, the start of phase 5 for the greater-region epidemic, i.e., the inflection point for *I*(*t*), seems to have already happened some time within the last 7-10 days (April 4-7th).

Overall, it seems that Greece is a few days ‘behind’ the progress of the COVID-19 outbreak in the greater region around it, with some countries being ‘ahead’ and others lagging ‘behind’ the average. It is extremely important to track these phase differences in the national epidemics, especially for countries that are closely interconnected via adjacent land borders, tourist shipping routes or direct flights. Without very strict border checking, large phase differences may translate to introduction of undetected infectious ‘seeds’ to a still non-immunized population, at least until an effective large-scale vaccination against COVID-19 becomes readily available in all countries.

## 7. Conclusion

COVID-19 constitutes a fast-pacing, world-wide pandemic that has evolved quickly into a multi-aspect international crisis. Even with proper policies and mitigation measures properly and promptly in place, tracking the outbreak even at the national level is an extremely challenging data analytics & modelling task as the event itself is still active, thus only limited and perhaps unreliable data are currently available.

In this study, Greece is the main focus for assessing the national outbreak and estimating the general trends and outlook of it. Multiple data analytics procedures, spectral decomposition and curve-fitting formulations are developed based on the data available at hand. Standard SIEQRDP epidemic modelling is applied for Greece and for the general region around it, providing hints for the outbreak progression in the mid- and long-term, for various infections under-reporting rates.

The overall short-term outlook for Greece seems to be towards positive, given the start of a downward trend in infections rate daily increase, a possible peak within a few days beyond April 14th, as well as the high availability level of ICU w.r.t. expected demand at peak. On the negative side, the fade-out period seems to be in the order of several months, with high probability of recurrent surges of the outbreak. The mitigation policies for the ‘next day’ should be focused on close tracking of the epidemic via large-scale tests, strict border checking in international travelling and an adaptive plan for selective activation of mitigation measures when deemed necessary.

## Data Availability

Data are already available by other open-access sources (John Hopkins CSSE).

## ACKNOWLEDGMENTS

The author wishes to thank every team and researcher that is currently working towards scientific works of high quality, readily accessible to the community, including technical papers, research papers and datasets. This is the only viable way to address such fast-pacing events of major world-wide impact collectively and effectively, as quickly and reliably as possible, while the crisis is still evolving.

## Footnotes

* [V. Wood, ‘Coronavirus 10 times more deadly than swine flu, says WHO’, The Independent (UK), 14-Apr-2020](https://www.independent.co.uk/news/health/coronavirus-deaths-cases-latest-swine-flu-who-a9462896.html).

† https://github.com/CSSEGISandData/COVID-19

‡ An inflection point of a function is where its concavity changes from upwards to downwards (‘falling’) or vice versa (‘rising’); more formally, the points where its curvature or second derivative changes sign, passing through zero-value in between.

§ https://imperialcollegelondon.github.io/covid19estimates/

¶ Estimated true Infected *≈* 0.13%(0.09%, 0.19%)*N*_*pop*_ of the general population or *I*(*t*) *≈* 13942(9652, 20377) w.r.t. reference demographics (2018) of *N*_*pop*_ = 10724599 (ELSTAT).

